# The Patients’ Voice in *Clostridioides difficile* Infection: Large Language Model-Assisted Thematic Analysis of Patient Testimonials

**DOI:** 10.64898/2026.07.08.26357545

**Authors:** Javier A. Villafuerte-Gálvez, Marco A. Noriega, Sena Cakir Colak, Carl V. Crawford

**Affiliations:** Division of Gastroenterology, Beth Israel Deaconess Medical Center, Harvard Medical School, Boston, MA, USA; Department of Medicine, Mount Auburn Hospital, Harvard Medical School, Cambridge, MA, USA; Division of Gastroenterology and Hepatology, Weill Cornell Medicine, New York, NY, USA

**Keywords:** *Clostridioides difficile* infection, patient-reported outcomes, large language models, thematic analysis, qualitative research, recurrent infection, fecal microbiota transplantation, caregiver burden

## Abstract

**Background:** *Clostridioides difficile* infection (CDI) imposes a burden that extends well beyond the gastrointestinal tract, yet existing outcome measures only partially capture the patient experience. We used frontier large language models (LLMs) on patient and caregiver narratives at scale to describe how burden shifts with disease course.

**Methods:** We analyzed 189 testimonials from the Peggy Lillis Foundation corpus, sorted into four cohorts with recurrence (*r*) and fulminant (*f*) severity as axes (rfCDI, fCDI, rCDI, non-rfCDI). Two independent LLMs coded eight thematic domains, four fulminant flags, thirteen emerging semantic fields, the dominant dimension, and narrative arcs. Two clinicians independently coded a subset for inter-rater reliability (PABAK, Gwet’s AC1).

**Results:** Treatment trajectory was the dominant theme in recurrent disease, whereas death and near-death dominated non-recurrent fulminant narratives. Psychological burden was near-universal in fulminant disease (98.0% in rfCDI, 97.2% in fCDI). Caregiver and bereavement content concentrated in fCDI (66.7%). Diagnostic failure was frequent across recurrent cohorts (47.6 - 56.1%). Bacteriotherapy tracked recurrence (60.2% rfCDI versus 5.6% fCDI). Financial, mental-health, and caregiver burdens were prominent and are currently unaddressed by guidelines. Human-human reliability was substantial (PABAK 0.79 for semantic fields, 0.76 for domains); arc coding was least reliable.

**Conclusions:** Patient narratives reveal a course-dependent, multidimensional burden in CDI. Concrete gaps exist between what patients prioritize, what guidelines recommend, and what therapy access provides. Frontier-LLM coding, validated against clinicians, offers a reproducible route to translate these priorities into research, care, and policy.

**Key Points:** Patient narratives reveal a course-dependent CDI burden: recurrence organizes around treatment, fulminant disease around survival, and bereavement. Psychological, financial, and caregiver domains important to patients remain largely unaddressed by current guidelines.

## 1. Introduction

*Clostridioides difficile* infection (CDI) remains one of the most common and consequential healthcare-associated and community-onset infections in the United States [1–3]. National surveillance data suggest a decline in incidence over the past decade, inviting optimism [1], though this warrants caution. Falling case counts partly reflect diagnostic stewardship and adoption of two-step nucleic acid amplification test (NAAT)-plus-toxin algorithms, driven by avoidance of financial penalties, rather than a true decline in disease burden [4,5]. Clinically important outcomes nonetheless persist, as recurrent CDI (rCDI) complicates a substantial proportion of index episodes [6], and mortality in fulminant CDI (fCDI) remains above 30% [7,8]. Access to advanced therapeutics has narrowed: Bezlotoxumab, the only approved monoclonal antibody for rCDI prevention, was withdrawn from the US market in early 2025 [9,10], and fecal microbiota transplantation (FMT) is now available only through structured investigational programs, following tighter oversight after exceedingly rare serious adverse events [11,12]. Newer live biotherapeutic products (LBPs) are effective for rCDI but lack efficacy for fCDI and carry considerable cost and sometimes limited formulary coverage, restricting their use [13].

The Cdiff32 is the leading validated CDI-specific patient-reported outcome instrument, measuring physical, mental, and social quality-of-life domains [14]. It has since been incorporated as a secondary outcome in trials of both approved LBPs, fecal microbiota spores, live-brpk (Vowst®), and fecal microbiota, live-jslm (Rebyota®), where active therapy was associated with meaningful improvements [15,16]. Its principal strength is the mental domain, sensitive to the anxiety, emotional distress, and fear of recurrence recognized as fundamental to the rCDI experience [17]. Structured instruments, however, constrain what patients may want to convey by design, capturing predefined domains that may miss critical parts of the experience we observe in clinical practice.

Direct qualitative studies of the lived experience of CDI outside clinical trials exist but remain scarce [18], and the weight accorded to patient and advocate perspectives in CDI practice guidelines has shifted across eras and is unevenly reflected in guideline-development processes [19]. Regulators have at times solicited patient and stakeholder input, and public comment was central to the advisory and approval process for LBPs [20]; nevertheless, broad access to FMT remains curtailed [11,12]. This misalignment between policy and the patients, caregivers, and clinicians increases the value of amplifying patients’ voices.

Patient-advocacy organizations such as the Peggy Lillis Foundation (PLF) have assembled large collections of patient testimonials, an underused resource for understanding the lived experience of CDI. Rigorous qualitative thematic analysis is labor-and resource-intensive, and scaling it to a corpus of hundreds of narratives was recently impossible for independent investigators, limiting the breadth of qualitative CDI research. Frontier large language models (LLMs) now make this feasible, provided the approach is conducted with methodological rigor [21,22]. In this context, we sought to characterize the PLF CDI testimonial corpus by clinical phenotype and narrative content, to apply a structured, LLM-assisted thematic coding framework across testimonials from individuals with CDI, and to assess the reliability of that coding through systematic comparison with independent human coders.

## 2. Methods

### 2.1 Study Design, Ethics, and Data Source

We conducted a retrospective thematic analysis of publicly posted CDI testimonials submitted by patients and caregivers to the PLF for advocacy and education. Because it used only public material and involved no interaction with individuals, it did not qualify as human-subjects research under 45 CFR 46.102, and institutional review board approval was not required; no identifying information is reported, and quotations are limited to non-identifying excerpts. The corpus of 204 narratives was retrieved from cdiff.org with a retrieval script (Supplemental Methods §S0.1).

### 2.2 Cohort Classification

We classified each testimonial using a three-step protocol, summarized in Supplemental Figure S3: a regular-expression pre-screen, independent dual-LLM classification (Claude Sonnet 4.6, DeepSeek V4 Pro) by recurrence, illness severity (including fulminant criteria[23]), and authorship, and concordance-based or PI adjudication of disagreements; full definitions and the adjudication protocol appear in Supplemental Methods §S0.2-S0.4. The PI adjudicated 34 discordant items. Recurrence and severity defined four analytic cohorts.

### 2.3 Thematic Coding Framework

We coded all testimonials against three sets of dimensions. Eight a priori domains (D1–D8) were applied to every cohort: diagnostic journey (D1), treatment trajectory (D2), physical symptom burden (D3), psychological and emotional burden (D4), social and functional disruption (D5), healthcare-system experience (D6), information-seeking and patient agency (D7), and recovery and resilience (D8). Four fulminant-specific flags (F1–F4) applied only to the fulminant cohorts (rfCDI and fCDI): sepsis or organ failure (F1), intensive care or ventilation (F2), emergency surgery (F3), and death or near-death (F4). Frequent emergent expressions not captured by the D and F items were identified by the primary LLM coder (Claude Sonnet 4.6) and clustered semantically into 13 emergent semantic fields (SF01–SF13) by the principal investigator (J.V.G); these were then coded de novo across the whole corpus. Each dimension and semantic field was coded as present or absent. We also coded the single domain that best characterizes a testimonial’s primary focus as the dominant domain, and one narrative arc from a 14-category typology based on Frank’s illness-narrative archetypes[24]. The full definitions, the dominant-domain rubric, and the arc typology appear in the Supplemental Methods.

### 2.4 LLM Coding and Prompt Calibration

Both models were queried at temperature zero to maximize determinism. A structured calibration preceded the full-corpus run, using 60 synthetic testimonials generated from themes in published qualitative CDI literature[25–27] to tune the coding prompt against a ground truth independent of the model’s output (Supplemental Methods §S1.2). The most parsimonious effective prompt added explicit coding thresholds for the adjacent domains D5, D6, and D7 (Claude macro-F1 0.717 to 0.792; DeepSeek 0.847 to 0.855) and generalized to a held-out validation set; we deployed it unchanged. For each dimension within each cohort, we averaged the two models’ codings for the frequency analyses and retained the individual codings for the reliability analyses.

### 2.5 Inter-Rater Reliability

Two senior internal medicine residents, blinded to each other and to all model output, independently coded a 16-testimonial subsample (IRR-001 to IRR-016) selected to span the four cohorts and the framework’s edge cases. After primary coding, a calibration session reviewed the dimensions, revealing substantial disagreement; each rater then independently recoded seven testimonials. The principal investigator adjudicated 28 coding disagreements across these seven testimonials to produce the reference standard. We report three comparisons: between the two coders (human–human, n=16), between each coder and Claude Sonnet 4.6 (human–LLM, n=16), and between the two models (LLM–LLM, n=189). The prevalence- and bias-adjusted kappa (PABAK) was the primary metric [28], and Gwet’s AC1 is reported for convergent validity [29]; Cohen’s kappa [30] is shown unless the absolute difference from PABAK exceeds 0.30, indicating a prevalence paradox, which is flagged. PABAK values were interpreted using the Landis and Koch thresholds [31].

### 2.6 Statistical Analysis

We report each dimension’s frequency as the percentage of the relevant cohort in which the dimension was present. Frequencies for the fulminant flags F1–F4 are expressed relative to the combined fulminant cohort (rfCDI + fCDI; n=67). We performed no inferential testing on corpus frequencies since the PLF corpus constitutes the full study sample, and the objective is characterization rather than inference. Analyses were performed in Python 3.13.

### 2.7 Reproducibility and Use of AI

All model prompts, the synthetic calibration corpus, the analysis code, and the per-testimonial coding outputs are available (Data Availability). Claude models were used to assist with drafting, revision, condensation, and integrity checking of the manuscript. Grammarly was used for spelling and grammar review. The investigators designed the study, defined all coding criteria, adjudicated all discordant classifications, and take full responsibility for the data, its interpretation, and the manuscript.

## 3. Results

### 3.1 Corpus and Analytic Sample

We retrieved 204 CDI testimonials from the PLF corpus. Fifteen were excluded because neither model could confidently classify recurrence or illness severity, leaving 189 testimonials for analysis (Figure 1). These fell into four cohorts: recurrent fulminant CDI (rfCDI; n=49), fulminant CDI without reported recurrence (fCDI; n=18), recurrent non-fulminant CDI (rCDI; n=84), and single-episode non-fulminant CDI (non-rfCDI; n=38). Patient demographics, authorship, and acquisition characteristics are summarized in Table 1, Figure 2, and Supplemental Figure S2.

**Figure 1.**
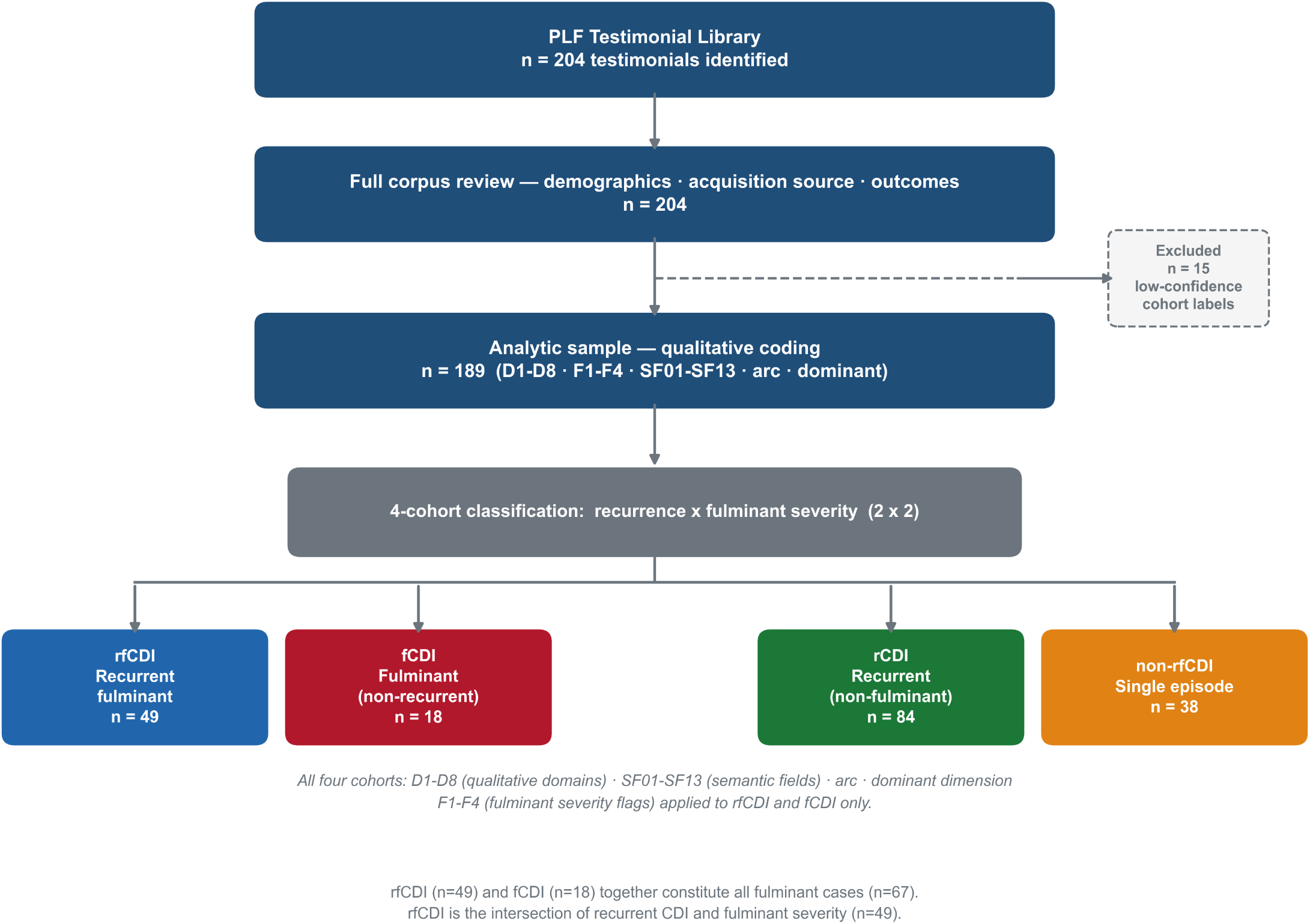
Study flow - PLF CDI testimonial corpus.

**Figure 2.**
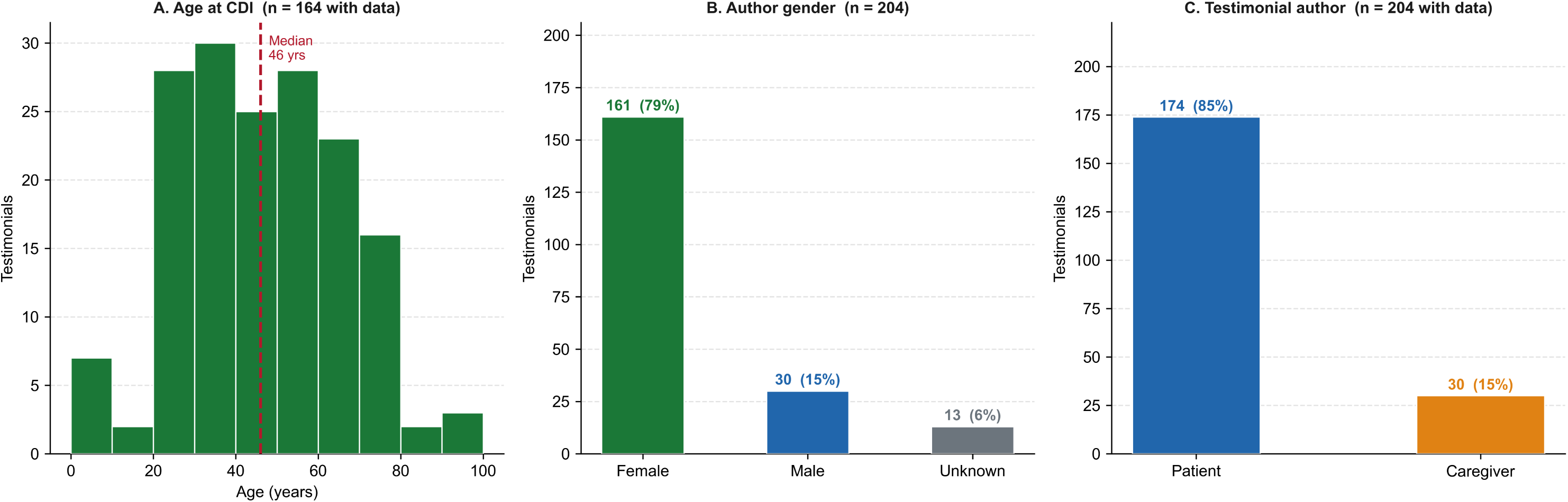
Demographic characteristics of PLF testimonial authors (n = 204)

**Table 1.** Sample characteristics by CDI cohort.

| Characteristic | rfCDI<br>(n=49) | fCDI<br>(n=18) | rCDI<br>(n=84) | non-rfCDI<br>(n=38) | Total<br>(n=189) | CDI<br>Reference<sup>b</sup> |
| --- | --- | --- | --- | --- | --- | --- |
| **n** | 49 | 18 | 84 | 38 | 189 | 615<sup>b</sup> |
| Female sex, n (%) | 45/49<br>(91.8%) | 12/18<br>(66.7%) | 67/84<br>(79.8%) | 28/38<br>(73.7%) | 152/189<br>(80.4%) | 329/615 (53.3%)<sup>b</sup> |
| Age, mean (range)<sup>a</sup> | 46.1 (5–<br>92) | 58.4 (14–<br>87) | 40.8 (1–<br>91) | 51.1 (24–<br>77) | 45.9 (1–<br>92) | Median 68<br>(IQR 55–77)<sup>b</sup> |
| Caregiver-authored, n (%) | 8/49<br>(16.3%) | 12/18<br>(66.7%) | 10/84<br>(11.9%) | 0/38<br>(0.0%) | 30/189<br>(15.9%) | — |
| FMT/bacteriotherapy<br>mentioned, n (%) | 27/49<br>(55.1%) | 1/18<br>(5.6%) | 35/84<br>(41.7%) | 4/38<br>(10.5%) | 67/189<br>(35.4%) | — |
| Illness duration, median<br>months [IQR] | 7 [3.5–18] | 3 [2–7] | 4 [1.5–12] | 2.5 [1–6] | 4 [1.2–12] | — |

<sup>a</sup> Age data available for: rfCDI n=38, fCDI n=15, rCDI n=70, non-rfCDI n=31.
<sup>b</sup> CDI reference cohort: Alonso CD et al. *Clin Infect Dis.* 2022;74:2142-2149 (PMID 34537841); n=615 hospitalized CDI patients at two US academic medical centers. Age and sex from Table 1 of that publication (verified from PDF). Note: community-associated CDI skews younger (median age 50–51 vs. ~72 for healthcare-associated CDI; Fu Y et al. *Ther Adv Gastroenterol.* 2021, PMID 34093740). National CDI incidence is higher among females (rate ratio 1.26 vs. males) and adults ≥65 years (rate ratio 8.65 vs. younger; Lessa FC et al. *N Engl J Med.* 2015;372:825-834, PMID 25714160). Caregiver-authored, FMT-mentioned, and illness duration rows are specific to PLF testimonial-based data collection and have no published population comparator.
fCDI, fulminant CDI (non-recurrent); rfCDI, recurrent fulminant CDI; rCDI, recurrent non-fulminant CDI; non-rfCDI, non-recurrent non-fulminant CDI; FMT, fecal microbiota transplantation; IQR, interquartile range.

### 3.2 Thematic Domains Across Cohorts (D1-D8)

Two domains approached universality across all cohorts: treatment trajectory (D2) and physical symptom burden (D3). The remaining domains followed a severity gradient: psychological/emotional burden (D4) and healthcare-system experience (D6) concentrated in the sicker cohorts, while recovery and outlook (D8) ran in the opposite direction (Table 2, Panel A; Figure 3).

**Figure 3.**
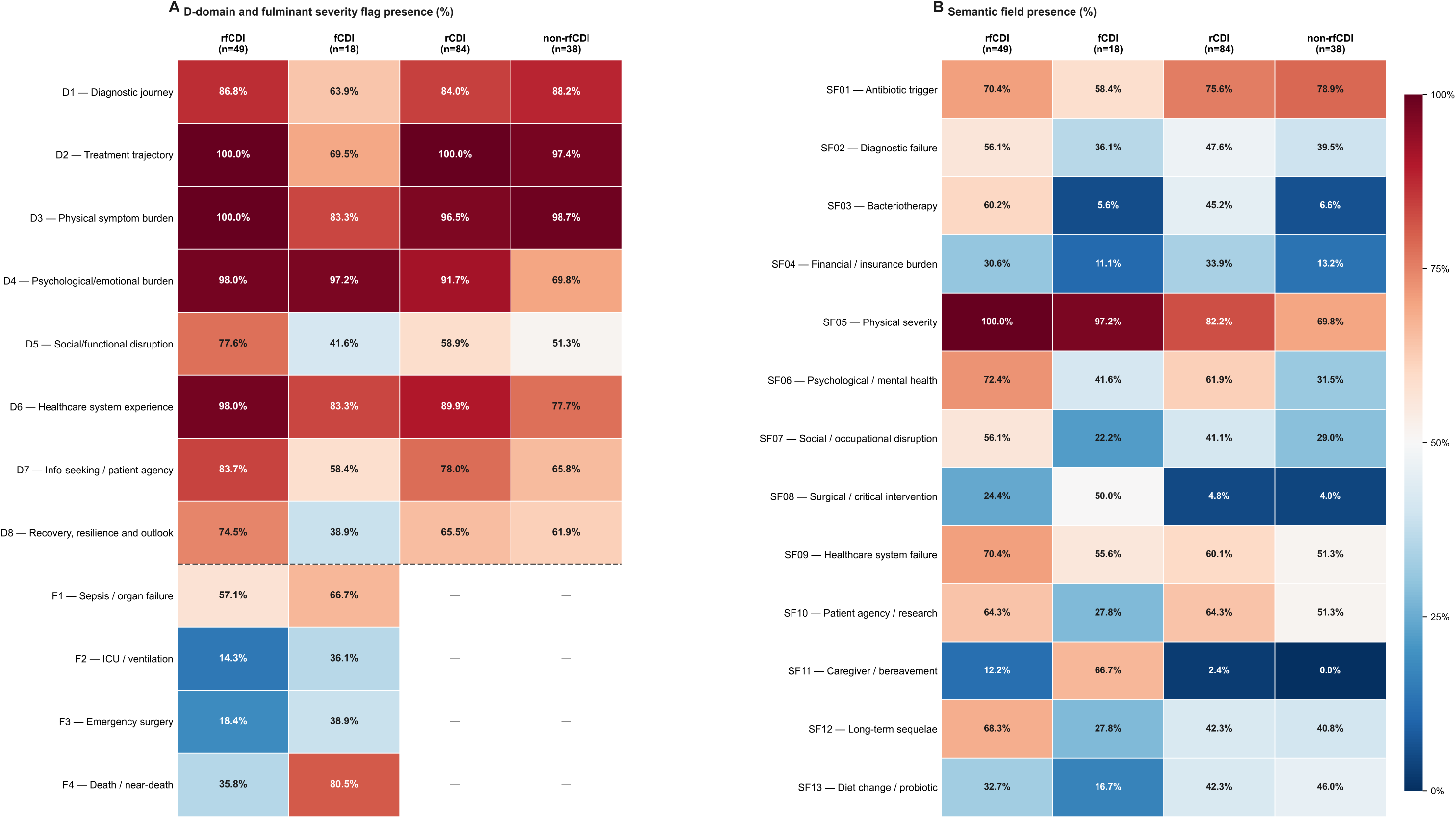
Demographic characteristics of PLF testimonial authors (n = 204)

**Table 2.** Thematic coding domain, fulminant severity flag, and semantic field presence (%) by CDI cohort.

**Panel A. Thematic coding domains (D1–D8) and fulminant severity flags (F1–F4)**
|  | rfCDI<br>(n=49) | fCDI<br>(n=18) | rCDI<br>(n=84) | non-rfCDI<br>(n=38) |
| --- | --- | --- | --- | --- |
| D1 — Diagnostic journey | 86.8% | 63.9% | 84.0% | 88.2% |
| D2 — Treatment trajectory | 100.0% | 69.5% | 100.0% | 97.4% |
| D3 — Physical symptom burden | 100.0% | 83.3% | 96.5% | 98.7% |
| D4 — Psychological/emotional burden | 98.0% | 97.2% | 91.7% | 69.8% |
| D5 — Social/functional disruption | 77.6% | 41.6% | 58.9% | 51.3% |
| D6 — Healthcare system experience | 98.0% | 83.3% | 89.9% | 77.7% |
| D7 — Info-seeking / patient agency | 83.7% | 58.4% | 78.0% | 65.8% |
| D8 — Recovery, resilience and outlook | 74.5% | 38.9% | 65.5% | 61.9% |
| F1 — Sepsis / organ failure | 57.1% | 66.7% | — | — |
| F2 — ICU / ventilation | 14.3% | 36.1% | — | — |
| F3 — Emergency surgery | 18.4% | 38.9% | — | — |
| F4 — Death / near-death | 35.8% | 80.5% | — | — |

*Note: Fulminant severity flags (F1–F4) apply only to fulminant cohorts; rCDI and non-rfCDI cells are not applicable (—). Emergency surgery (F3, 38.9%) exceeds bacteriotherapy in fCDI (SF03, 5.6%), indicating patients reached surgical intervention before microbiome-based therapy was accessible.*

**Panel B. Semantic fields (SF01–SF13)**
|  | rfCDI<br>(n=49) | fCDI<br>(n=18) | rCDI<br>(n=84) | non-rfCDI<br>(n=38) |
| --- | --- | --- | --- | --- |
| SF01 — Antibiotic trigger | 70.4% | 58.4% | 75.6% | 78.9% |
| SF02 — Diagnostic failure | 56.1% | 36.1% | 47.6% | 39.5% |
| SF03 — Bacteriotherapy | 60.2% | 5.6% | 45.2% | 6.6% |
| SF04 — Financial / insurance burden | 30.6% | 11.1% | 33.9% | 13.2% |
| SF05 — Physical severity | 100.0% | 97.2% | 82.2% | 69.8% |
| SF06 — Psychological / mental health | 72.4% | 41.6% | 61.9% | 31.5% |
| SF07 — Social / occupational disruption | 56.1% | 22.2% | 41.1% | 29.0% |
| SF08 — Surgical / critical intervention | 24.4% | 50.0% | 4.8% | 4.0% |
| SF09 — Healthcare system failure | 70.4% | 55.6% | 60.1% | 51.3% |
| SF10 — Patient agency / research | 64.3% | 27.8% | 64.3% | 51.3% |
| SF11 — Caregiver / bereavement | 12.2% | 66.7% | 2.4% | 0.0% |
| SF12 — Long-term sequelae | 68.3% | 27.8% | 42.3% | 40.8% |
| SF13 — Diet change / probiotic | 32.7% | 16.7% | 42.3% | 46.0% |

*Abbreviations: CDI, Clostridioides difficile infection; fCDI, fulminant CDI (non-recurrent); rfCDI, recurrent fulminant CDI; rCDI, recurrent non-fulminant CDI; non-rfCDI, non-recurrent non-fulminant CDI; FMT, fecal microbiota transplantation; D1–D8, qualitative coding domains; F1–F4, fulminant severity flags; SF01–SF13, semantic fields.*

Treatment trajectory (D2) was the modal dominant dimension overall (Figure 4); the two models diverged most in the rfCDI cohort, where DeepSeek assigned D4 as the primary domain far more often than Claude (30.6% versus 6.1%, section 3.6).

**Figure 4.**
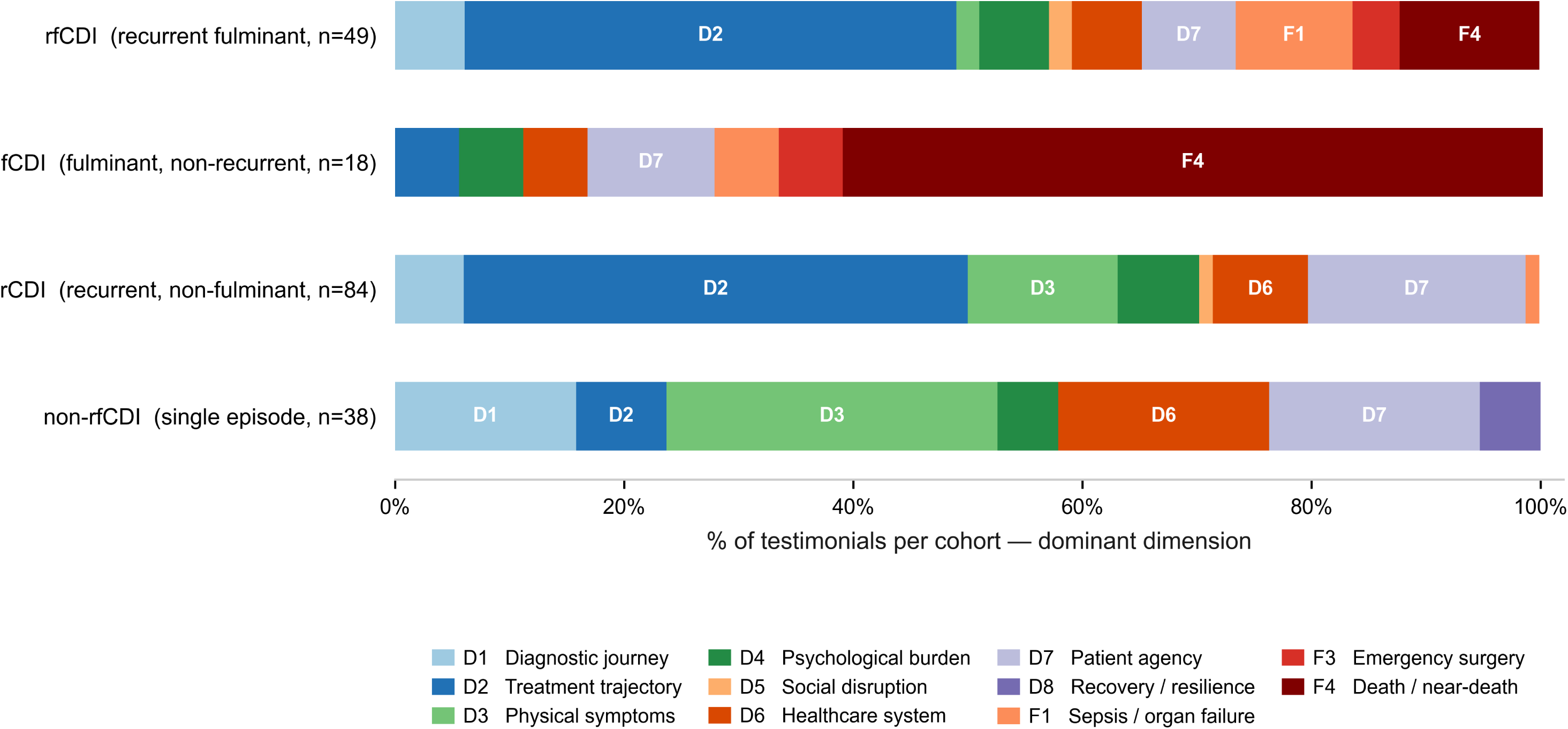
Primary thematic frame (dominant dimension) by CDI cohort (Claude Sonnet 4.6 codings)

### 3.3 Fulminant-Specific Flags (F1-F4)

The fulminant flags apply only to the fulminant cohorts (n=67). Death or near-death (F4) dominated the fCDI cohort, consistent with its larger share of caregiver-authored narratives, whereas intensive care/ventilation (F2) was the least frequent flag in both fulminant cohorts; full counts are shown in Table 2 (Panel A).

### 3.4 Semantic Fields (SF01-SF13)

Antibiotic exposure (SF01) was evenly distributed across cohorts, while physical severity language (SF05) was nearly universal in fulminant cohorts and high elsewhere (Table 2, Panel B). The starkest contrast was in caregiver and bereavement content (SF11), concentrated mainly in fCDI. Psychological/mental-health content (SF06) was graded by severity, and surgical or critical intervention (SF08) clustered in the fulminant cohorts.

Healthcare-system failure (SF09) and diagnostic failure (SF02) were common across all groups. Financial/insurance burden (SF04) was more frequent in recurrent cases, and access to bacteriotherapy (SF03) closely followed recurrence, remaining low in fCDI and non-rfCDI. Figure 5 summarizes the key qualitative features of the four cohorts, with examples in Supplemental Table S5.

**Figure 5.**
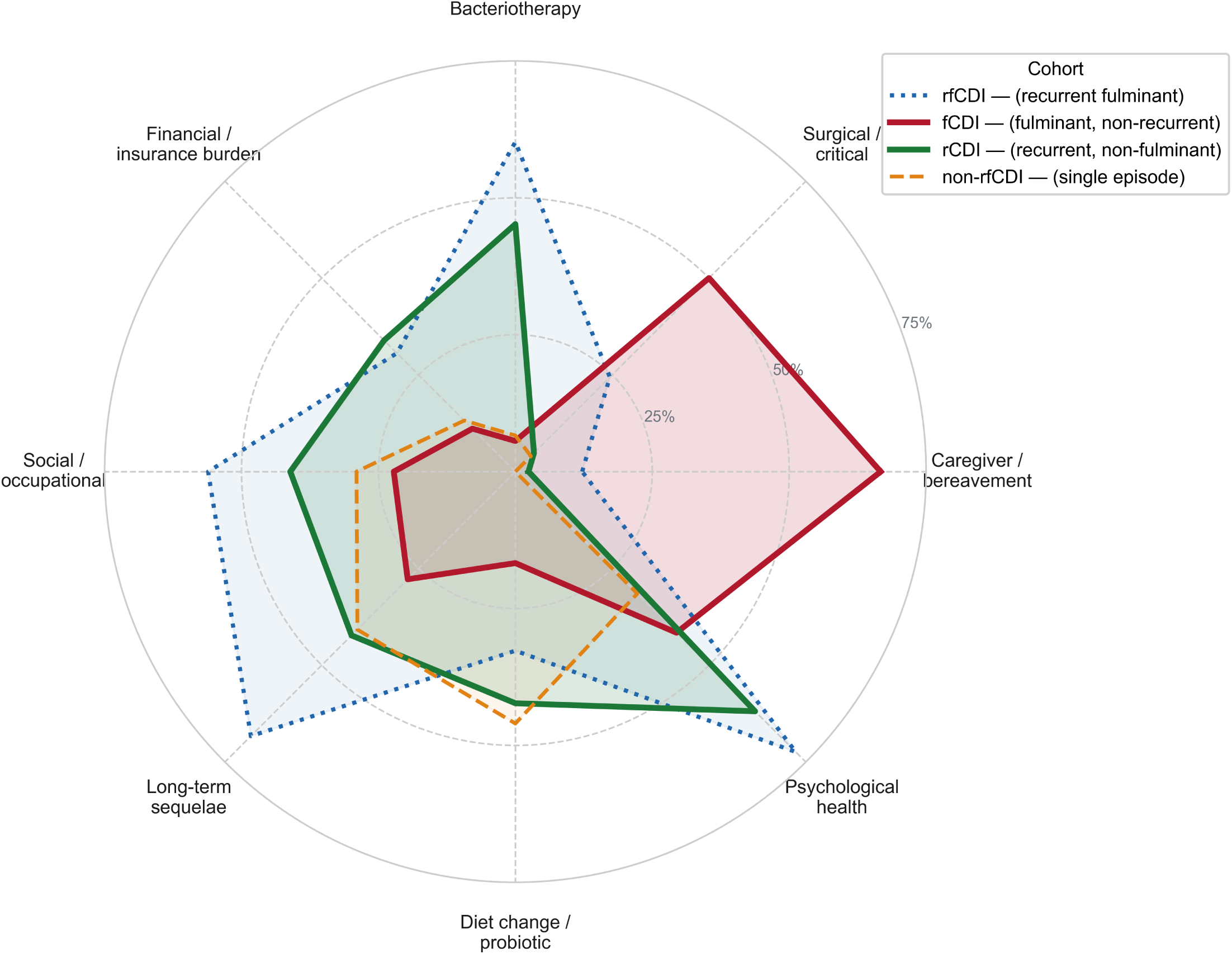
Top 8 discriminating features by CDI cohort (D-domains + semantic fields; ranked by coefficient of variation; % of testimonials)

### 3.5 Narrative Arcs (Exploratory)

The narrative arc assignment is treated as exploratory throughout, as this layer showed the lowest inter-rater agreement (section 3.6). With that caveat, the fCDI cohort was notably concentrated in a caregiver-burden arc, consistent with its high SF11 frequencies, while the rfCDI cohort was more distributed across resolution-via-FMT, near-death-to-recovery, and caregiver-burden arcs. The rCDI and non-rfCDI cohorts showed the most diffuse distributions, with the two models diverging substantially on individual arcs; full distributions by cohort appear in Supplemental Figure S1.

### 3.6 Coding Reliability

Human-human PABAK averaged 0.788 across the 13 semantic fields and 0.757 across the 12 binary D and F dimensions, while the percent agreement for arc assignment was 68.8%. These values establish a hierarchy of agreement: semantic fields were coded most consistently, binary domains at an intermediate level, and arc assignment least reliably, which supports the exploratory designation of the arc layer (Supplemental Table S3).

Human-LLM agreement on the semantic fields (PABAK 0.596-0.731) fell below the human-human benchmark, with consistent directional patterns. Human coders applied SF06 (psychological/mental health), SF07 (social/occupational disruption), and SF10 (patient agency/research) more liberally, whereas the models applied SF02 (diagnostic failure) more liberally than either human rater. Full agreement statistics, including a comparison of LLM-LLM and human-human reproducibility, appear in Supplemental Table S4.

## 4. Discussion

This study of 189 patient and caregiver narratives that spanned the full spectrum of *Clostridioides difficile* infection (CDI) revealed that patients experience CDI not as a self-limited enteric infection but as a chronic, multidimensional illness. The physical symptoms the medical community focuses on are only one component of a much broader ordeal encompassing medical, psychological, social, and financial burdens. This burden was also not uniform across different manifestations of CDI [32].

The dominant frame of each narrative shifted systematically with disease phenotype: recurrent disease organized itself around treatment trajectory, the dominant dimension in 44.0% of rCDI testimonials, whereas non-recurrent fulminant disease organized itself around death and near-death, dominant in 61.1% of the fCDI cohort (Figure 4). Symptoms of CDI thus appear to evolve across the disease course rather than representing a single, static entity, an evolution the analysis that follows treats as its central object.

Full domain, flag, and semantic-field frequencies by cohort are shown in Table 2 and Figure 3 (Panel A: D1–D8/F1–F4; Panel B: SF01–SF13). For patients with rCDI, repeated antibiotic courses, serial physician encounters, recurrent stool testing, insurance barriers, and failed therapies convert what physicians have always believed to be an acute infection into a chronic medical condition (D2, coded in 100.0% of recurrent narratives). Rather than discrete episodes of diarrhea, patients described months to years of uncertainty in which every improvement was shadowed by the fear of relapse – a phenomenology in which microbiological cure repeatedly failed to translate into restoration of ordinary life. This distinction matters clinically since physicians typically gauge success by decreased recurrence rates or return to normal stool form, whereas patients gauge it by restoration of daily function, confidence in future health, and freedom from constant vigilance, factors that shape long-term plans, employment, and even the use of medically necessary antibiotics. Recurrence, from the patient’s perspective, is less a mere repeated infection and more a chronic illness state with psychosocial consequences that extend beyond the gastrointestinal tract.

On the other hand, those experiencing more life-threatening forms of CDI, namely fCDI, inverted this experience. Fulminant narratives centered on survival itself, given the high rates of sepsis, intensive care unit needs, emergency colectomy, and near-death or death. A striking feature of this cohort was authorship: caregiver and bereavement-focused content (SF11) was far more common in fCDI than in any other cohort, and nearly two-thirds of fCDI testimonials were written by family members rather than by patients (Table 1). This reflects an unavoidable and unfortunate selection, since many patients with catastrophic CDI succumb before narrating their own illness, and their experience reaches us only through a bereaved relative who becomes an advocate, decision maker, and witness. The burden of fulminant disease, unfortunately, thus extends beyond the patient to caregivers who have been largely absent from prior qualitative CDI research. Future outcome instruments and survivorship studies should incorporate caregiver perspectives to fully characterize the severe CDI experience.

Although CDI is recognized for its debilitating physical manifestations, psychological distress was among the most consistent and dominant findings across the entire corpus. Emotional burden (D4, SF06) was near-universal in fulminant disease and remained common even in the non-fulminant narrative. This distress was not merely a reaction to severe diarrhea as narratives described an amalgam of recurrent relapse, prolonged uncertainty, isolation, fear of infecting loved ones, and eroded trust in clinicians, with the anxiety about future antibiotic exposure and relapse that often persist long after apparent clinical recovery. Unlike standardized questionnaires that quantify anxiety at a single point in time, these narratives illuminate how such morbidity develops longitudinally alongside repeated healthcare encounters and therapeutic failure. This is an integral component of the disease process that interventions aimed solely at eradicating CDI will not address. Routine assessment of anxiety, depression, and social functioning warrants greater consideration during longitudinal follow-up.

One of the most actionable findings was the prevalence of healthcare-system failure across all phenotypes (D6, SF09). Patients describe delayed diagnosis, fragmented care, and barriers to appropriate therapy. These accounts rarely portray a single grave error but rather a sequence of individually modest failures that accumulate over time. Diagnostic failure was especially prominent in rCDI (SF02), notable given that contemporary diagnostic assays possess excellent analytical performance. The rate-limiting step is not laboratory technology but clinical recognition. Patients were often reassured that symptoms represented irritable bowel syndrome (IBS), anxiety, or medication effects before repeat testing confirmed recurrence; rCDI is, it seems, more of a longitudinal diagnostic challenge that requires clinicians to revisit prior assumptions as symptoms evolve, rather than anchor to an initial impression. With this background, the degree of patient self-advocacy was more than expected (SF10). Patients independently consulted the primary literature, identified FMT, and appealed insurance denials. Patient agency seems to have developed as an adaptive response to prolonged illness amid perceived system deficiencies; the degree of self-navigation documented in the corpus is, unfortunately, better read as a marker of system failure than a hallmark of optimal patient-centered care.

Nearly half of all testimonials described long-term sequelae extending for months to years beyond microbiological cure, most frequently in rfCDI (SF12). These include chronic gastrointestinal symptoms, dietary restriction, fatigue, and durable impairment in quality of life. This “survivorship” phase, likely reflecting some combination of persistent microbiome disruption, post-infectious gut-brain axis disorders, and psychological trauma, is largely overlooked by conventional outcome measures. Many patients described an uninterrupted “CDI experience” and continue to identify as “having C. diff” despite repeated negative stool tests. Defining the biological and psychosocial determinants of CDI survivorship and testing interventions aimed at long-term recovery rather than recurrence prevention alone is a clear priority.

Self-directed lifestyle modification, including dietary restriction, probiotic use, and efforts to protect the microbiome, was common across all cohorts (SF13) and reinforced the concept of CDI as a chronic adaptive illness rather than an acute diarrheal one; many patients sought out probiotics despite guidelines concluding the evidence is insufficient for recurrence prevention, likely because these measures were among the few safe, accessible, and tangible actions available to reduce uncertainty about prognosis. Microbiome-directed therapy proper (FMT or other bacteriotherapy) showed a sharper contrast, far more common in recurrent than in fulminant or single-episode disease (SF03), with many patients describing it as the turning point that separated prolonged illness from recovery. Its near-absence from fulminant narratives reflects the historical period of many testimonials, evolving regulatory pathways, and these patients’ critical instability. That the therapies patients describe as decisive are becoming harder to reach, given bezlotoxumab’s withdrawal [10] and the narrowing of FMT access, even as guidelines conditionally endorse it for refractory severe or fulminant disease [33], warrants continued study as newer live biotherapeutic products become available.

Financial toxicity was a clinically under-recognized dimension of burden. Roughly a third of patients with recurrent disease described insurance denials, prolonged disability, lost employment, or substantial out-of-pocket costs (SF04), interacting with illness, hospitalization, and caregiving to amplify distress and delay recovery.

This was one of several recurring priorities, alongside psychological and mental health concerns, social and occupational disruptions, long-term sequelae, and caregiver burden, that share a common feature. There has been little explicit attention to these aspects in current societal guidance, which is organized mainly around diagnosis, antimicrobial selection, and infection prevention rather than downstream consequences. Mapping the patient-reported semantic fields against five major guideline documents (Table 3) localizes the gap. Each of these domains appears in one-to-two-thirds of fCDI narratives, yet is largely or wholly unaddressed across guidelines from infectious disease and gastroenterology societies [19,33–35]. This comparison is interpretative and post hoc since the corpus was not assembled to audit the guidelines. Table 3, shows the consistency of the omission, suggests that patient-centered domains could be incorporated into the patient-facing sections of future CDI guidance.

**Table 3.**
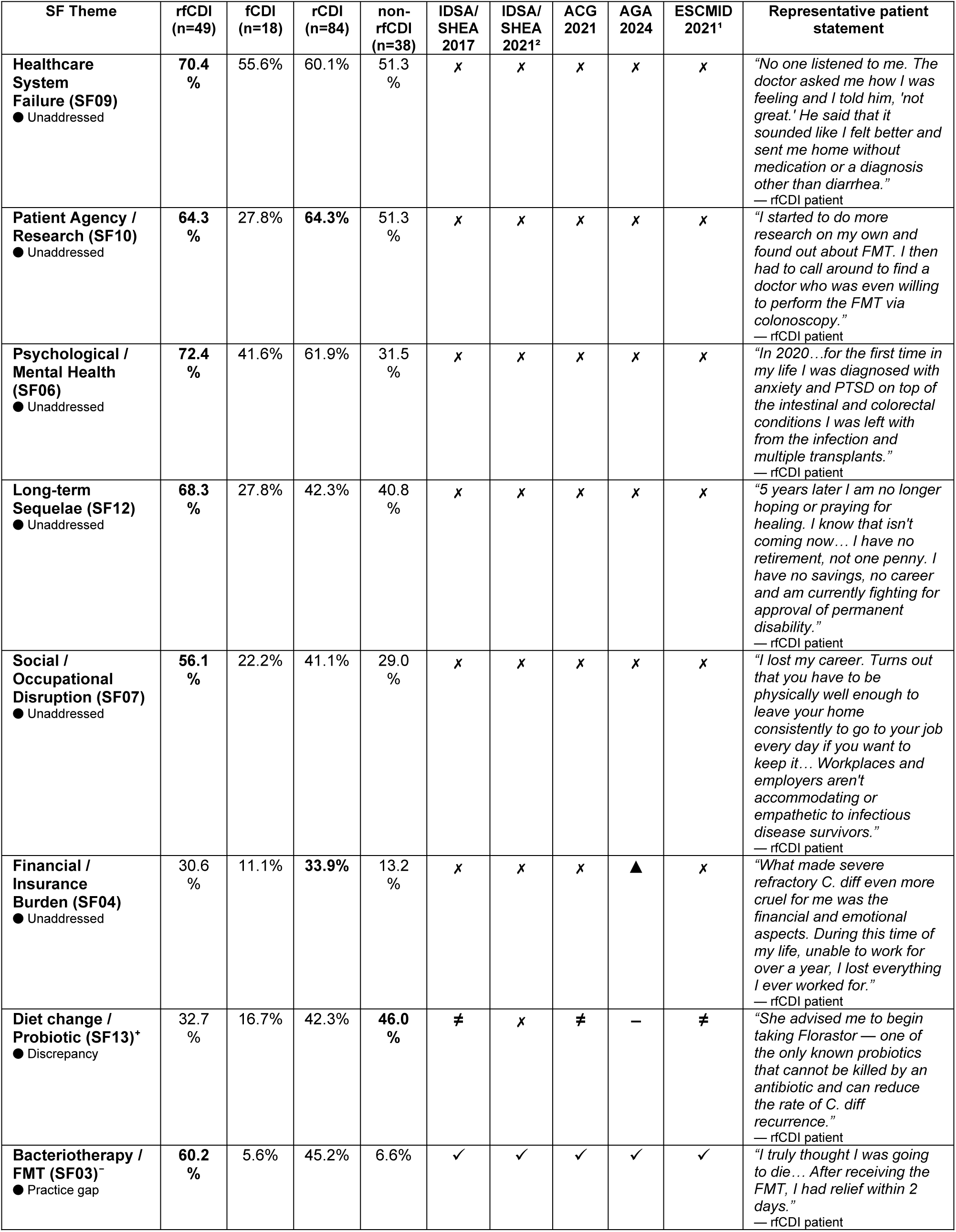
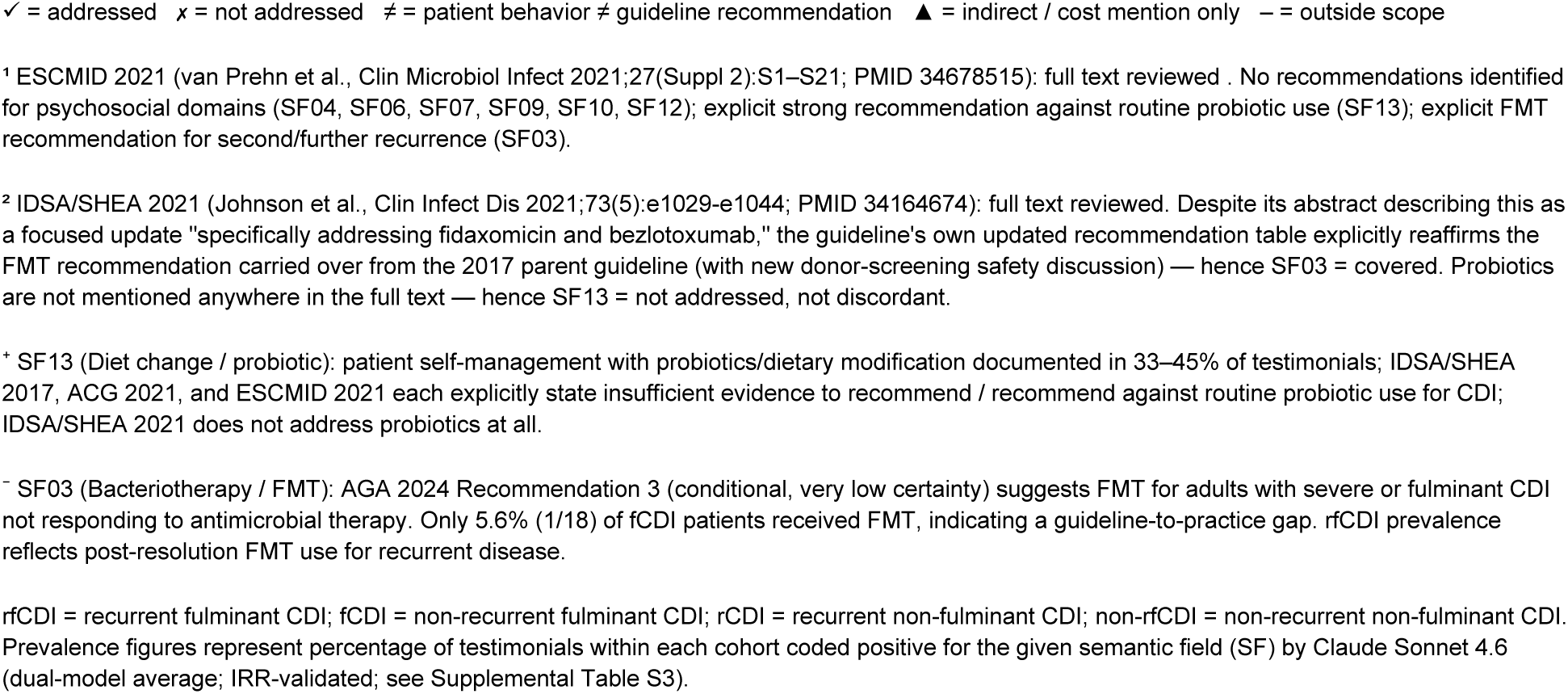
Patient-Reported Needs vs. Clinical Guideline Coverage in CDI.

| SF Theme | rfCDI<br>(n=49) | fCDI<br>(n=18) | rCDI<br>(n=84) | non-rfCDI<br>(n=38) | IDSA/<br>SHEA<br>2017 | IDSA/<br>SHEA<br>2021<sup>2</sup> | ACG<br>2021 | AGA<br>2024 | ESCMID<br>2021<sup>1</sup> | Representative patient<br>statement |
| --- | --- | --- | --- | --- | --- | --- | --- | --- | --- | --- |
| **Healthcare**<br>**System**<br>**Failure (SF09)**<br>● Unaddressed | **70.4**<br>**%** | 55.6% | 60.1% | 51.3<br>% | x | x | x | x | x | "No one listened to me. The doctor asked me how I was feeling and I told him, 'not great.' He said that it sounded like I felt better and sent me home without medication or a diagnosis other than diarrhea."<br>— rfCDI patient |
| **Patient Agency /**<br>**Research (SF10)**<br>● Unaddressed | **64.3**<br>**%** | 27.8% | **64.3%** | 51.3<br>% | x | x | x | x | x | "I started to do more research on my own and found out about FMT. I then had to call around to find a doctor who was even willing to perform the FMT via colonoscopy."<br>— rfCDI patient |
| **Psychological /**<br>**Mental Health**<br>**(SF06)**<br>● Unaddressed | **72.4**<br>**%** | 41.6% | 61.9% | 31.5<br>% | x | x | x | x | x | "In 2020...for the first time in my life I was diagnosed with anxiety and PTSD on top of the intestinal and colorectal conditions I was left with from the infection and multiple transplants."<br>— rfCDI patient |
| **Long-term**<br>**Sequelae (SF12)**<br>● Unaddressed | **68.3**<br>**%** | 27.8% | 42.3% | 40.8<br>% | x | x | x | x | x | "5 years later I am no longer hoping or praying for healing. I know that isn't coming now... I have no retirement, not one penny. I have no savings, no career and am currently fighting for approval of permanent disability."<br>— rfCDI patient |
| **Social /**<br>**Occupational**<br>**Disruption (SF07)**<br>● Unaddressed | **56.1**<br>**%** | 22.2% | 41.1% | 29.0<br>% | x | x | x | x | x | "I lost my career. Turns out that you have to be physically well enough to leave your home consistently to go to your job every day if you want to keep it... Workplaces and employers aren't accommodating or empathetic to infectious disease survivors."<br>— rfCDI patient |
| **Financial /**<br>**Insurance**<br>**Burden (SF04)**<br>● Unaddressed | 30.6<br>% | 11.1% | **33.9%** | 13.2<br>% | x | x | x | ▲ | x | "What made severe refractory C. diff even more cruel for me was the financial and emotional aspects. During this time of my life, unable to work for over a year, I lost everything I ever worked for."<br>— rfCDI patient |
| **Diet change /**<br>**Probiotic (SF13)\***<br>● Discrepancy | 32.7<br>% | 16.7% | 42.3% | **46.0**<br>**%** | ≠ | x | ≠ | — | ≠ | "She advised me to begin taking Florastor — one of the only known probiotics that cannot be killed by an antibiotic and can reduce the rate of C. diff recurrence."<br>— rfCDI patient |
| **Bacteriotherapy /**<br>**FMT (SF03)<sup>-</sup>**<br>● Practice gap | **60.2**<br>**%** | 5.6% | 45.2% | 6.6% | ✓ | ✓ | ✓ | ✓ | ✓ | "I truly thought I was going to die... After receiving the FMT, I had relief within 2 days."<br>— rfCDI patient |

✓ = addressed ✕ = not addressed ≠ = patient behavior ≠ guideline recommendation ▲ = indirect / cost mention only – = outside scope
<sup>1</sup> ESCMID 2021 (van Prehn et al., Clin Microbiol Infect 2021;27(Suppl 2):S1–S21; PMID 34678515): full text reviewed . No recommendations identified for psychosocial domains (SF04, SF06, SF07, SF09, SF10, SF12); explicit strong recommendation against routine probiotic use (SF13); explicit FMT recommendation for second/further recurrence (SF03).
<sup>2</sup> IDSA/SHEA 2021 (Johnson et al., Clin Infect Dis 2021;73(5):e1029-e1044; PMID 34164674): full text reviewed. Despite its abstract describing this as a focused update "specifically addressing fidaxomicin and bezlotoxumab," the guideline's own updated recommendation table explicitly reaffirms the FMT recommendation carried over from the 2017 parent guideline (with new donor-screening safety discussion) — hence SF03 = covered. Probiotics are not mentioned anywhere in the full text — hence SF13 = not addressed, not discordant.
<sup>+</sup> SF13 (Diet change / probiotic): patient self-management with probiotics/dietary modification documented in 33–45% of testimonials; IDSA/SHEA 2017, ACG 2021, and ESCMID 2021 each explicitly state insufficient evidence to recommend / recommend against routine probiotic use for CDI; IDSA/SHEA 2021 does not address probiotics at all.
<sup>~</sup> SF03 (Bacteriotherapy / FMT): AGA 2024 Recommendation 3 (conditional, very low certainty) suggests FMT for adults with severe or fulminant CDI not responding to antimicrobial therapy. Only 5.6% (1/18) of fCDI patients received FMT, indicating a guideline-to-practice gap. rfCDI prevalence reflects post-resolution FMT use for recurrent disease.

Reassuringly, the themes that surface spontaneously in these narratives align closely with the three subscales of the Cdiff32, supporting its construct validity with patient-generated data, while also surfacing four further process-of-care domains (diagnostic journey, treatment trajectory, healthcare-system experience, and patient agency) without a dedicated Cdiff32 subscale [14,36] (Table 2).

To our knowledge, this is among the first studies applying frontier LLMs to thematic coding of an entire patient-narrative corpus validated against an independent human inter-rater study, which showed models and clinicians were differentially reproducible: clinicians on interpretive, context-dependent judgments and models on threshold-defined and holistic ones, with the exploratory arc layer least reliable for both [21,22,37] (Supplemental Table S4).

There are several limitations to this study. This design is descriptive by intent and involves no inferential testing, so cohort contrasts should be read as patterns rather than as claims of significance. The corpus is a self-selected advocacy sample from a single English-language platform, enriched for severe, prolonged, and treatment-refractory disease. The prevalence figures describe this population of motivated narrators rather than CDI experience at large, consistent with our study population skewing younger and more frequently female than a well-characterized CDI cohort from the Northeastern US [23]. Clinical details were patient- or caregiver-reported and unverified against records, and recurrence followed a patient-reported definition rather than the clinical 8- to 12-week criterion, a deliberate choice to honor how patients mark a new episode but one that limits nosological precision. Thematic coding itself carries limitations as some findings may reflect model behavior rather than unmediated patient testimony, most notably the illness arc layer, which had the least reliable coding in our inter-rater study (human-human agreement 68.8%; human-LLM 37.5% to 43.8%) and is accordingly reported as exploratory throughout. The same inter-rater work showed systematic human-LLM divergence on interpretative semantic fields such as psychological/mental health and social/occupational disruption, raising the possibility that model coding under-detects implied psychosocial content, though this may partly reflect coder expertise rather than true LLM underperformance.

The construction of this thematic code for CDI, despite its limitations, points toward a novel research direction that treats patient priorities as a measurement target rather than dismissing them. Pairing narrative corpora with validated instruments such as Cdiff32 in prospective cohorts could test whether the domains patients raised most, within financial strain, caregiver burden, and mental health, predict downstream outcomes that current measures miss [17,38]. Extending beyond a single advocacy platform to multi-source, non-English corpora would test how far these patterns generalize. The overarching message is that patients define recovery differently from clinicians – physicians appropriately prioritize eradicating infection, preventing recurrence, and avoiding complications, and guidelines reflect those priorities; patients value these too, but their narratives make it clear that recovery also encompasses the restoration of daily functioning, confidence in future health, financial stability, social participation, and trust in the healthcare system. Until these dimensions are routinely incorporated into clinical research, quality-improvement work, patient-reported outcome measures, and future guidelines, an essential component of the CDI experience will remain unmeasured and, too often, unaddressed.

## Acknowledgments

Code: Python/matplotlib visualization scripts were drafted and refined with Claude Sonnet 4.6 assistance and verified in detail by the authors against the underlying data.

Images: The visual abstract and supplemental methods figures were drafted as JSON-structured prompts, refined iteratively with Claude Sonnet 4.6 assistance, and generated with the nano-banana-pro-preview model via the Google AI Studio API (google-genai client); all were reviewed and verified by the authors.

Language editing: Grammar and language were checked with Grammarly; all edits were reviewed and accepted by the authors.

The authors thank the Peggy Lillis Foundation and, above all, the patients and families whose narratives made this work possible.

## Data Availability

De-identified coding outputs, model prompts, analysis code, the synthetic calibration corpus, and the coding dictionary are openly available at https://doi.org/10.5281/zenodo.21262959. The underlying testimonial narratives are publicly posted by their authors via the Peggy Lillis Foundation and are available from the Foundation on reasonable request; they are not redistributed here to protect the identifiability of the narrators.

## Ethics Statement

This study analyzed patient and caregiver narratives that were voluntarily and publicly posted by their authors through the Peggy Lillis Foundation. The investigators determined that this activity does not meet the definition of human subjects research under the U.S. Department of Health and Human Services’ Common Rule (45 CFR 46.102(e)(1)): the narratives were not obtained through intervention or interaction with the narrators, and they do not constitute private information, as the authors posted them voluntarily for a public audience.

## Author Contributions

J.V.G.: conceptualization, methodology, formal analysis, visualization, data curation, writing - original draft, supervision. M.A.N. and S.C.C: investigation (blinded independent inter-rater coding), methodology (IRR design input), writing - review & editing. C.V.C.: conceptualization, writing – original draft (Discussion), writing - review & editing.

