## Supplemental Methods, Tables, Figures for "The Patients’ Voice in *Clostridioides difficile* Infection: Large Language Model-Assisted Thematic Analysis of Patient Testimonials"

### Supplementary Methods

*LLM-assisted coding of CDI patient testimonials: full technical methods*

#### Contents

S0. Cohort Classification Protocol
S1. LLM Coding Pipeline
S2. Coding Framework: D/F Domain Definitions and SF01-SF13 Definitions
S3. Narrative-Arc Typology and Dominant-Domain Rubric
S4. Synthetic Benchmark Design and Ground-Truth Independence
S5. Three-Iteration Prompt Tuning
S6. Held-Out Validation
S7. Inter-rater Reliability Protocol
References

*Table SM1. Thematic Domain Framework (D1-D8; F1-F4)*
*Table SM2. Narrative-Arc Typology and Frank (1995) Archetype Mapping*
*Table SM3. Three-Iteration Prompt Tuning Results*
*Table SM4. Held-Out Validation GO/NO-GO Decision*
*Table SM5. Per-Domain Validation F1 (n=20)*

#### Abbreviations and Notation

| Abbreviation | Meaning |
|---|---|
| **CDI** | Clostridioides difficile infection |
| **rCDI** | recurrent CDI |
| **fCDI** | fulminant CDI, non-recurrent |
| **rfCDI** | recurrent and fulminant CDI |
| **non-rfCDI** | single-episode, non-fulminant CDI |
| **FMT** | fecal microbiota transplantation |
| **ICU** | intensive care unit |
| **LLM** | large language model |
| **PABAK** | prevalence-adjusted bias-adjusted kappa |
| **AC1** | Gwet's first-order agreement coefficient |
| **kappa (k)** | Cohen's kappa |
| **IRR** | inter-rater reliability |
| **SF** | semantic field (SF01-SF13) |

| | |
|---|---|
| **D/F** | thematic domain codes (D1-D8 standard; F1-F4 fulminant-specific) |
| **n** | number of testimonials |
| **temperature** | LLM sampling temperature (0 = deterministic) |
| **JSON** | JavaScript Object Notation (structured text output format) |

### S0. Cohort Classification Protocol

#### S0.1 Corpus Assembly

The Peggy Lillis Foundation (PLF) testimonial corpus comprised 204 publicly posted CDI narratives, retrieved from cdiff.org in May 2026 with a Python HTML-parsing pipeline. Each narrative was stored as structured text (the testimonial_en field of the structured corpus file; available from the Foundation on reasonable request) under a unique identifier. When one narrative appeared under more than one identifier, the duplicate was resolved in favor of the fulminant-cohort record, so that severe presentations were not lost to deduplication. No narrative was excluded for length; the only exclusions were the 15 low-confidence items defined in S0.5.

#### S0.2 Regex Pre-Screen

A deterministic keyword screen flagged narratives that explicitly mentioned fulminant-compatible events (for example, ICU, colectomy, sepsis, ventilator, ileostomy, colostomy, or toxic megacolon). The screen was a triage step only: flagged narratives were prioritized for fulminant review, and every narrative was classified by both models regardless of the screen result. The screening keywords are defined in the scraping script (plf_extract.py), a sanitized copy of which is provided in the replication package.

#### S0.3 Dual-LLM Classification

Each narrative was independently classified by two large language models, Claude Sonnet 4.6 and DeepSeek V4 Pro, using the system prompt and user template reproduced below (reclassify_all_cohorts.py in the replication package). Both models were queried at a temperature of 0, the setting that makes the output deterministic: the same narrative returns the same classification on every run. The prompt elicited three judgments per narrative: recurrence (yes, no, or unclear), severity (uncomplicated, severe, fulminant, or unclear), and authorship (patient or caregiver).

##### System Prompt (verbatim)

```
You are a clinical reviewer classifying CDI (Clostridioides difficile
infection) patient testimonials. Respond with valid JSON only -- no prose
before or after.


Classify on THREE dimensions:


--- DIMENSION 1: RECURRENCE ---
Did the author (or the patient they describe) experience MORE THAN ONE CDI
episode?
Definition: any new CDI episode after a prior one resolved or was treated, per
the text.
Patient self-report is sufficient. Time interval does NOT matter.
Fields:
  "recurrent": "yes" | "no" | "unclear"
  "recurrence_evidence": direct quote <=2 sentences supporting your call, or ""
```

```
  "episode_count_implied": "1" | "2" | "3+" | "unclear"

--- DIMENSION 2: SEVERITY (most severe CDI episode described) ---
Fields:
  "severity": "uncomplicated" | "severe" | "fulminant" | "unclear"
  "severity_criteria_present": array (strings from fulminant list below, may be
empty)
  "severity_evidence": direct quote <=2 sentences supporting severity call, or
""

Definitions:
  uncomplicated -- outpatient management, oral antibiotics, no hospitalization
required
  severe        -- hospitalization required; significant symptoms; NO fulminant
criteria
  fulminant     -- ANY of the following directly attributed to CDI:
    icu_admission          mechanical_ventilation      bowel_surgery_cdi
    sepsis_septic_shock    hypotension                 elevated_wbc
    acute_kidney_injury    tachycardia_sirs            toxic_megacolon
    death_from_cdi         near_death                  organ_failure

  Patient-reported language counts for fulminant criteria -- examples:
    "very high white count" -> elevated_wbc
    "kidneys failed" / "renal failure" -> acute_kidney_injury
    "heart rate in the 160s" / "my heart was racing at 160" -> tachycardia_sirs
    "blood pressure crashed" / "went into shock" -> hypotension
    Do not require medical terminology; flag the clinical equivalent.

--- DIMENSION 3: AUTHORSHIP ---
  "written_by": "patient" | "caregiver" | "unclear"

--- CONFIDENCE & NOTES ---
  "confidence": "high" | "medium" | "low"
  "notes": any concern warranting PI attention, or ""

Example response:
{
  "recurrent": "yes",
  "recurrence_evidence": "I had three separate bouts of C. diff over two
years",
  "episode_count_implied": "3+",
  "severity": "fulminant",
  "severity_criteria_present": ["icu_admission", "sepsis_septic_shock"],
  "severity_evidence": "I was admitted to the ICU and developed sepsis
throughout my body",
  "written_by": "patient",
  "confidence": "high",
  "notes": ""
}
```

#### User Message Template (verbatim)

```
Testimonial slug: {slug}
```

```
{text}
```

{slug} was replaced with the testimonial identifier; {text} was replaced with the full narrative text from the structured corpus file, testimonial_en field. Recurrence was defined broadly as any patient-reported new CDI episode after prior resolution, without requiring the clinical 8- to 12-week interval criterion, which is rarely documented in lay testimonials; time interval did not affect classification (see System Prompt, Dimension 1).

#### S0.4 Concordance and PI Adjudication

When both models returned the same recurrence and severity labels with high confidence, the item was assigned by consensus (n=125); these concordant classifications were further verified with a 10-item concordant spot-check before being finalized. When the models disagreed on a single dimension, a prespecified rule assigned the label from the model that had proven more accurate for that dimension during calibration: recurrence from Claude Sonnet 4.6, severity from DeepSeek V4 Pro (rule-applied, n=45). Items that disagreed on both dimensions or returned "unclear" were adjudicated by the principal investigator, who reviewed the full narrative and both model outputs before assigning a final label (PI-adjudicated, n=34). Fulminant attribution followed the standard of Alonso and colleagues [1], counting a qualifying event (ICU admission, colectomy, or death) only when it was primarily or substantially attributable to CDI rather than to a comorbid condition.

#### S0.5 Low-Confidence Exclusions

Fifteen narratives whose recurrence or severity remained unclear after each classification step were designated as low confidence and excluded from the analytic corpus. These exclusions reflect classification ambiguity, not brevity: the 15 narratives range from 52 to 596 words (median 209). They remain in the master corpus file but contribute to no frequency or reliability analysis.

#### S0.6 Analytic Cohorts

Narratives with non-unclear recurrence and severity labels were assigned to one of four mutually exclusive cohorts: rfCDI (recurrent and fulminant; n=49), fCDI (fulminant, non-recurrent; n=18), rCDI (recurrent, non-fulminant; n=84), and non-rfCDI (single-episode, non-fulminant; n=38). The analytic corpus totaled 189. The fulminant denominator (rfCDI plus fCDI) is 67, and the recurrent denominator (rfCDI plus rCDI) is 133. Unless a cohort subset is specified, all frequency analyses use n=189.

### S1. LLM Coding Pipeline

#### S1.1 Models and Settings

Primary coder: Claude Sonnet 4.6 (Anthropic; model ID claude-sonnet-4-6; accessed May 2026). Independent second coder: DeepSeek Chat (DeepSeek AI; model ID deepseek-chat, corresponding to DeepSeek V4 Pro; accessed May 2026). Both models were called at temperature 0 for the full-corpus coding run, ensuring deterministic outputs within each model run. The Anthropic Messages API and DeepSeek OpenAI-compatible Chat Completions API were used directly via Python urllib without third-party wrappers. The full-corpus coding script is run_full_dataset.py in the replication package.

#### S1.2 System Prompt and D5/D6/D7 Calibration Addendum

The tuned iteration-1 system prompt (baseline plus the D5/D6/D7 calibration addendum) was used for both models in the full-corpus run (n=189). The prompt consisted of: (1) role framing and task description; (2) standard domain definitions (D1-D8); (3) fulminant-specific domain definitions (F1-F4, coded null for rCDI and non-rfCDI items); (4) narrative arc typology (14 arc labels); (5) dominant domain instruction; and (6) a threshold calibration addendum for domains D5, D6, and D7, which were the primary source of inter-model disagreement in baseline testing. The calibration addendum is reproduced verbatim below. The prompt was tuned using 60 synthetic testimonials generated from themes in three published qualitative studies of CDI and related gastrointestinal conditions, so that ground-truth domain labels derived from the source literature rather than from any model output; the synthetic corpus was divided into a 40-item tuning set and a 20-item validation set, and three prompt iterations were evaluated on the tuning set, maximizing the macro-averaged F1 across D1-D8, before the selected prompt was generalized to the validation set and deployed unchanged.

**D5/D6/D7 Calibration Addendum (verbatim, embedded in production prompt):**

```
## Threshold Calibration for Adjacent Domains (D5, D6, D7)

These three domains are frequently confused. Apply the following rules
strictly:

**D5 -- Social/functional disruption**
Code 1 ONLY if the testimonial explicitly describes: (a) missing work, being
unable to work, losing a job, or going on disability; OR (b) withdrawing from
social activities, canceling plans, or becoming homebound due to CDI; OR (c)
relationship strain explicitly caused by the illness.
Code 0 if symptoms are described but no explicit social or functional
consequence is mentioned. Feeling ill is NOT enough -- the disruption must be
named.

**D6 -- Healthcare system experience**
Code 1 if the testimonial describes meaningful friction or interaction with the
healthcare system: dismissive clinicians, diagnostic delays by clinicians,
insurance denial, difficulty getting referrals, or poor care coordination.
```

```
Code 0 if the patient merely mentions going to a doctor or receiving treatment
without describing a barrier, inadequacy, or systemic issue.


**D7 -- Information seeking & patient agency**
Code 1 ONLY if the patient (or caregiver) describes active agency: researching
CDI independently, seeking a second opinion, finding a specialist themselves,
advocating for a specific test or treatment, or joining support communities.
Code 0 if the patient passively followed their doctor's recommendations, even
if they received good information or a novel treatment.
```

#### S1.3 Response Format and Parsing

Each model was prompted to return a single JSON object containing binary domain flags (D1-D8: 0 or 1; F1-F4: 0, 1, or null for rCDI and non-rfCDI items), a narrative-arc label, and a dominant-domain label. Claude Sonnet 4.6 occasionally wrapped fulminant testimonial responses in markdown code fences (```json...```); the parser stripped these before json.loads(). DeepSeek was called with response_format={'type': 'json_object'}, which enforced bare JSON output. Both parsers included error handling with per-item failure logging; failed items were recorded with error metadata and excluded from aggregate metrics.

#### S1.4 Caching and Resumability

Each API response was cached as a JSON file, keyed by the testimonial identifier, before any parsing occurred. If a cache file existed, the API was not re-called. This design allowed the full-corpus run to be safely interrupted and resumed without re-incurring API costs or introducing non-determinism from re-calls.

### S2. Coding Framework

#### S2.1 D/F Domain Definitions

The following domains were applied systematically across all coded testimonials (n=189 analytic corpus). Domains D1-D8 were applied to all four cohorts. Domains F1-F4 were applied to fulminant testimonials only (rfCDI n=49, fCDI n=18; F-flag denominator n=67). See Table SM1.

**Table SM1. Thematic Domain Framework (D1-D8; F1-F4)**

| Code | Domain Name | Definition | Applied to |
|---|---|---|---|
| **D1** | **Diagnostic journey** | Experiences related to initial CDI diagnosis: delays, misdiagnosis, route to correct diagnosis, atypical presentations, repeated negative tests. | All cohorts |
| **D2** | **Treatment trajectory** | Antibiotic sequences, treatment failures, FMT and biologic therapy (Vowst, Zinplava), treatment decision-making, cost of treatment. | All cohorts |
| **D3** | **Physical symptom burden** | Somatic experience of CDI: profuse diarrhea, abdominal pain, weight loss, dehydration, hair loss, reactive arthritis, secondary physical sequelae. | All cohorts |
| **D4** | **Psychological / emotional impact** | Anxiety, depression, PTSD, suicidal ideation, CDI hypervigilance, fear of recurrence, stigma, grief, loss of pre-CDI identity. | All cohorts |
| **D5** | **Social / functional disruption** | Impact on work and career, financial consequences, parenting disruption, loss of relationships, social isolation, disability. Code 1 ONLY if explicitly named; symptoms alone are not sufficient. | All cohorts |
| **D6** | **Healthcare system experience** | Insurance barriers, access to specialists, clinician communication failures, diagnostic errors, institutional harms, care coordination. Code 1 if meaningful friction is described. | All cohorts |
| **D7** | **Information-seeking & patient agency** | Self-directed research, demand for specific tests or treatments, dietary/probiotic self-management, escalation to specialists, PLF advocacy. Code 1 only if active agency, not passive receipt of care. | All cohorts |
| **D8** | **Recovery, resilience &** | Narrative of improvement, adaptation to ongoing sequelae, persistent physical or | All cohorts |

| | | | |
|---|---|---|---|
| | **outlook** | psychological effects after cure, forward-looking orientation. | |
| **F1** | **Sepsis / organ failure** | Sepsis or septic shock, hemodynamic instability requiring vasopressors, acute kidney injury, respiratory failure, coagulopathy (DIC), documented organ system failure. | Fulminant only (n=67) |
| **F2** | **ICU / life support** | ICU admission, mechanical ventilation, vasopressor support, renal replacement therapy, total parenteral nutrition, prolonged sedation. | Fulminant only (n=67) |
| **F3** | **Major surgical intervention** | Emergent or urgent colectomy, loop ileostomy, other major abdominal surgery directly attributed to CDI or its complications. | Fulminant only (n=67) |
| **F4** | **Death / near-death outcome** | Death directly attributed to CDI (caregiver narrative) or near-death experience: cardiac arrest, explicit clinician statement of imminent death, near-death survivor language. | Fulminant only (n=67) |

*CDI = Clostridioides difficile infection; rCDI = recurrent CDI; ICU = intensive care unit; DIC = disseminated intravascular coagulation; FMT = fecal microbiota transplantation. All coding was binary (0 = theme absent, 1 = theme present). A single dominant domain was assigned per testimonial. Calibration thresholds for D5, D6, and D7 (see S1.2) were added during prompt tuning. Cohort n's: rfCDI=49, fCDI=18, rCDI=84, non-rfCDI=38; total analytic n=189.*

#### S2.2 SF01-SF13 Semantic Field Definitions

Thirteen binary semantic fields (SF01-SF13) were applied to the full analytic corpus (n=189) by both LLM coders. Each field was coded 1 (present) or 0 (absent) per testimonial. Multiple fields can be positive within a single testimonial. Field boundaries were adjudicated by the PI on 2026-05-17. Full definitions follow; display labels for figures and tables use approved short forms (SF03 = 'Bacteriotherapy'; SF13 = 'Diet change / probiotic').

##### SF01 -- Antibiotic Trigger

**Definition:** Antibiotics are named or implied as the precipitating cause of CDI: prior antibiotic use disrupted the microbiome, enabling C. difficile colonization.

Include: patient explicitly states antibiotics caused or preceded CDI; specific triggering antibiotics named (e.g., fluoroquinolones, clindamycin, broad-spectrum penicillins/cephalosporins); hospitalization context where antibiotic use is strongly implied as trigger.
Exclude: antibiotics used only for CDI treatment (vancomycin, fidaxomicin); hospital exposure without any mention of prior antibiotics.
Borderline: 'I was on clindamycin for a dental procedure and got C. diff' -- SF01=1. 'I developed C. diff after my hip replacement surgery' -- code 1 only if antibiotic use is implied or stated. 'I used the hospital bathroom' -- no antibiotic trigger; SF01=0.

##### SF02 -- Diagnostic Failure

**Definition:** Delayed diagnosis, misdiagnosis, or clinician dismissal before the correct CDI identification was made.

Include: weeks or months elapsed between symptom onset and CDI diagnosis; prior diagnosis of another condition (IBS, anxiety, Crohn's) before CDI; emergency department or clinic visit in which CDI was not diagnosed; stool testing not ordered despite symptoms consistent with CDI; clinician dismissed or minimized symptoms.
Exclude: rapid or uncomplicated diagnosis (code 0); diagnostic tests that led promptly to correct diagnosis.
Borderline: 'It took 6 months to diagnose me' -- SF02=1. 'My doctor said it was just anxiety' -- SF02=1. 'I was diagnosed the same day I saw a doctor' -- SF02=0.

##### SF03 -- Bacteriotherapy

**Definition:** Use of or reference to fecal microbiota transplantation (FMT) or an FDA-approved microbiome-restoring biotherapeutic (fecal microbiota spores, live-brpk [Vowst®]; fecal microbiota, live-jslm [Rebyota®]).

Include: FMT (received, pursued, or discussed as option); Vowst® (fecal microbiota spores, live-brpk) mentioned; Rebyota® (fecal microbiota, live-jslm) mentioned; microbiome restoration described as a treatment approach.
Exclude: probiotics only (yogurt, Culturelle, etc.) without FMT or biotherapeutics; antibiotics used as CDI treatment (vancomycin, fidaxomicin).
Borderline: 'My doctor mentioned FMT as the next step' -- SF03=1. 'I had FMT from a donor stool' -- SF03=1. 'I took Culturelle daily' -- SF03=0.

##### SF04 -- Financial / Insurance Burden

**Definition:** Out-of-pocket costs, insurance denials, prior authorization hurdles, or affordability barriers to CDI treatment.

Include: insurance denial for a specific CDI treatment (FMT, Vowst®, Rebyota®, fidaxomicin); out-of-pocket costs explicitly mentioned; financial hardship due to CDI treatment costs; pharmacy or insurer blocking access to a treatment on cost grounds.
Exclude: work loss/income loss (code SF07 instead); general treatment access barriers without a financial dimension (code SF09).
Borderline: 'My insurance denied Vowst and I had to pay $18,000' -- SF04=1. 'The pharmacy said Dificid was too expensive' -- SF04=1. 'My insurance covered everything' -- SF04=0.

##### SF05 -- Physical Severity

**Definition:** Severe, debilitating, or life-threatening physical manifestations of CDI, beyond mild gastrointestinal upset.

Include: high-frequency diarrhea (profuse, uncontrollable, or severely disruptive); significant weight loss (any amount mentioned); hospitalization for CDI symptoms; dehydration, electrolyte imbalance requiring intervention; fatigue or weakness preventing normal activities; post-CDI physical sequelae: persistent diarrhea, IBS, reactive arthritis, hair loss.
Exclude: mild diarrhea described as manageable without hospitalization (use context); psychological symptoms only.
Borderline: 'I lost 40 pounds' -- SF05=1. 'I had 15 bouts of diarrhea per day' -- SF05=1. 'I had loose stools for a week' -- code based on severity described.

##### SF06 -- Psychological / Mental Health

**Definition:** Depression, anxiety, PTSD symptoms, suicidal ideation, grief, trauma, stigma, or other mental health impact attributable to CDI.

Include: anxiety, panic attacks, agoraphobia, fear of relapse; depression, hopelessness, despair; suicidal ideation or suicidal crisis explicitly described; stigma: feeling ashamed, dirty, or embarrassed about symptoms; PTSD-like symptoms, nightmares, hypervigilance post-CDI; grief: caregiver grief or patient grief over lost quality of life.
Exclude: social isolation alone (code SF07 if described without explicit emotional language); brief phrases like 'frustrated' or 'upset' without elaboration (borderline).
Borderline: 'I became deeply depressed' -- SF06=1. 'I was suicidal' -- SF06=1. 'I felt dirty and ashamed' -- SF06=1. 'I was frustrated' -- borderline; look for elaboration before coding 1.

##### SF07 -- Social / Occupational Disruption

**Definition:** Work loss, inability to fulfill social roles, social withdrawal, relationship strain, or loss of participation in activities of daily life.

Include: work absence, job loss, or inability to maintain employment; social withdrawal or avoiding social events due to CDI symptoms; relationship strain with partner, family, or friends; inability to care for children or other dependents; inability to participate in hobbies, travel, or prior lifestyle.
Exclude: psychological distress causing isolation (code SF06 as well); financial loss from medical costs (code SF04).
Borderline: 'I couldn't go to my daughter's wedding' -- SF07=1. 'I missed three months of work' -- SF07=1. 'I became a hermit' -- SF07=1.

##### SF08 -- Surgical / Critical Intervention

**Definition:** Emergency abdominal surgery directly caused by CDI (colectomy, ileostomy), or admission to ICU with mechanical ventilation or vasopressor support.

Include: total or subtotal colectomy; ileostomy creation (and reversal/takedown); emergency abdominal surgery described in CDI context; ICU admission for CDI (mechanical ventilation, vasopressors, life support); septic shock or hemodynamic instability described.
Exclude: diagnostic endoscopy or colonoscopy only; hospitalization on a general ward (not ICU).
Borderline: 'He had emergency surgery to remove his colon' -- SF08=1. 'She was on a ventilator in the ICU' -- SF08=1. 'I was hospitalized for a week' -- SF08=0 (no ICU/surgery described).

##### SF09 -- Healthcare System Failure

**Definition:** Inadequate clinician response, dismissal, poor coordination of care, or systemic barriers resulting in suboptimal CDI management.

Include: clinician dismissed or minimized symptoms; poor care coordination: inappropriate referrals, handoffs, communication gaps; inadequate information provided at diagnosis; emergency department visit with no treatment or incorrect treatment; system-level delays (e.g., prior authorization delays for biotherapeutics).
Exclude: insurance/financial barriers (code SF04); diagnostic delay (code SF02; may co-occur).
Borderline: 'My doctor never told me C. diff could recur' -- SF09=1. 'The ER sent me home with anti-nausea meds only' -- SF09=1. 'My gastroenterologist was excellent' -- SF09=0.

##### SF10 -- Patient Agency / Research

**Definition:** Active self-advocacy, independent research, self-referral to specialists, joining patient communities, or public advocacy.

Include: patient researched CDI online, in the medical literature, or via patient communities; self-referral to an infectious disease specialist, a tertiary center, or an FMT clinic; participation in PLF or other CDI patient organizations; writing letters, contacting legislators, speaking at events; educating clinicians or others about CDI.
Exclude: passive engagement (attending appointments) without initiative described; recovery without an active agency component (consider D8 arc).
Borderline: 'I found the PLF online and joined their support group' -- SF10=1. 'I demanded an ID referral' -- SF10=1. 'I went to my doctor when told to' -- SF10=0.

##### SF11 -- Caregiver / Bereavement

**Definition:** Testimonial written from a caregiver's perspective, OR describing an intensive caregiving burden, OR a patient's death described by a surviving family member.

Include: testimonial explicitly told by a spouse, parent, child, or other caregiver; author describes providing hands-on care (medication management, wound care, transport); describes a family member's CDI illness from a third-person perspective; patient died, and a caregiver/family member describes the loss.

Exclude: brief mention of family support without caregiving narrative; rCDI patient testimonials in first person about their own illness (unless also a caregiver for another person). Borderline: 'As told by his wife...' -- SF11=1. 'My husband cared for me during treatment' -- SF11=0 (brief support mention, not a caregiver narrative).

##### SF12 -- Long-term Sequelae

**Definition:** Persistent physical symptoms, functional impairment, or complications that continue after the acute CDI episode has resolved.

Include: persistent diarrhea or bowel dysfunction after CDI resolved; post-infectious IBS, food intolerances, reactive arthritis, hair loss; permanent functional impairment (permanent ileostomy, ongoing fatigue); long-term medication required to manage post-CDI bowel function; prolonged recovery described as lasting months after the infection. Exclude: ongoing active CDI (not yet resolved); psychological sequelae only (code SF06). Note: Distinguish from SF06 (psychological sequelae). A patient with both post-CDI IBS and PTSD is positive for both SF12 and SF06. Borderline: 'Even after treatment my bowels were never the same' -- SF12=1. 'I still have IBS two years later' -- SF12=1. 'I am still fighting C. diff' -- SF12=0 (active, unresolved).

##### SF13 -- Microbiome / Dietary Self-Management

**Definition:** Dietary modifications, probiotic supplementation, or other microbiome-focused self-management strategies used for CDI recovery or prevention.

Include: specific dietary changes described (low-fiber, low-sugar, BRAT diet, fermented foods); probiotic use (named brands or types: Saccharomyces boulardii, Lactobacillus, Culturelle, etc.); avoidance of trigger foods to prevent flares; self-experimentation with diet to manage bowel symptoms post-CDI. Exclude: FMT and biotherapeutics (code SF03 for those); general healthy eating mentioned without specific CDI/microbiome context. Borderline: 'I had to eliminate all sugar and alcohol for months' -- SF13=1. 'I took Culturelle daily' -- SF13=1. 'I tried to eat healthily' without specifics -- SF13=0.

### S3. Narrative-Arc Typology and Dominant-Domain Rubric

#### S3.1 Narrative-Arc Typology

Narrative-arc assignment drew on Frank's three illness-narrative archetypes [2], restitution, chaos, and quest, extended with CDI-specific variants, the original framework does not capture. The restitution arc describes narratives in which recovery is the expected outcome; the chaos arc, narratives without trajectory or resolution; the quest arc, narratives in which illness becomes a catalyst for meaning. Fourteen arc labels were operationalized for this corpus (Table SM2).

**Table SM2. Narrative-Arc Typology and Frank (1995) Archetype Mapping**

| Arc | Frank (1995) Archetype | Notes |
|---|---|---|
| **RESTITUTION ARCS** | | |
| resolution-via-FMT | Restitution | CDI-specific treatment variant; FMT or biotherapeutic is the narrative's central resolution event |
| self-directed-recovery | Restitution | CDI-specific; narrator's own research or advocacy drives recovery |
| chronic-sequelae | Restitution (partial) | CDI-specific; infection resolved but lasting physical or psychological sequelae persist |
| cautious-hope | Restitution (emerging) | CDI-specific; recovery underway but durability uncertain; hedging language present |
| **QUEST ARCS** | | |
| adversity-to-resilience | Quest | Matches Frank's quest archetype; illness fully resolved and framed as transformative |
| advocacy-focused | Quest | CDI-specific; illness as catalyst for awareness, policy reform, or systemic advocacy |
| near-death-to-recovery | Quest | CDI-specific (fulminant); survival of a life-threatening episode as the defining narrative event |
| **CHAOS ARCS** | | |
| ongoing-struggle | Chaos | Matches Frank's chaos archetype; CDI unresolved, narrator actively fighting but retains agency |

| | | |
|---|---|---|
| ongoing-desperate | Chaos | CDI-specific; extreme chaos with explicit loss of hope, exhaustion, or crisis language |
| treatment-odyssey | Chaos | CDI-specific; narrative organized around a sequence of multiple failed treatment attempts |
| delayed-diagnosis | Chaos | CDI-specific; significant diagnostic delay is the central narrative feature |
| caregiver-burden | Chaos | CDI-specific; caregiver or bereaved family member perspective; patient experience secondary |
| **CONTEXT-SPECIFIC ARCS** | | |
| postpartum-crisis | CDI-specific | CDI occurring in peripartum or postpartum period; intersection with pregnancy is a prominent narrative feature |
| early-onset | CDI-specific | CDI in a pediatric patient (<18) or young adult (<30); young age is a notable narrative feature |

*Frank AW. The Wounded Storyteller: Body, Illness, and Ethics. Chicago: University of Chicago Press; 1995. 'CDI-specific' indicates arcs extending beyond Frank's original three archetypes. Arc labels derived from this corpus; the 14-arc typology was synthesized by the primary LLM coder (Claude Sonnet 4.6) during inductive analysis.*

#### S3.2 Dominant-Domain Rubric

In addition to binary domain presence/absence (D1-D8), a single dominant domain was assigned to each testimonial: the domain that best characterizes the testimonial's primary focus, not merely its presence. The dominant domain rubric extended the binary coding prompt with the following decision rules:

- D2 vs. D4 disambiguation: D2 (treatment trajectory) is the dominant domain when the narrative is primarily organized around WHAT HAPPENED medically. D4 (psychological/emotional) is dominant when the narrative is primarily about HOW IT FELT: fear, depression, grief, emotional toll.
- D3 vs. D2/D4 disambiguation: D3 (physical symptom burden) is dominant only when the physical experience itself is the explicit primary subject. When symptoms are described as context for treatment decisions (D2) or emotional consequences (D4), those domains take precedence.
- D1 vs. D2 disambiguation: D1 (diagnostic journey) is dominant when misdiagnosis, diagnostic delay, or the route to diagnosis is the central story. D2 is dominant when the narrative has moved past diagnosis into the treatment sequence.
- D7 and D8: D7 (information-seeking/agency) is dominant only when the narrator's self-directed research or advocacy is explicitly the narrative's organizing spine. D8

(recovery/resilience) is dominant only when the narrative is primarily forward-looking: lessons learned, return to function.

Dominant domain results are reported in Supplementary Tables S3 and S4.

### S4. Synthetic Benchmark Design and Ground-Truth Independence

#### S4.1 Rationale

No open-access CDI qualitative dataset with individual-level human domain codings exists that could serve as a calibration target. A synthetic benchmark corpus was therefore generated using literature-grounded synthesis. The design preserved ground-truth independence from any LLM by deriving ground-truth labels from researcher-assigned themes in peer-reviewed qualitative studies, not from model outputs.

#### S4.2 Seed Literature

Three open-access qualitative studies of CDI and related gastrointestinal conditions were used as seeds [7,8,9]. Patient quotes and researcher-assigned theme labels from each paper were extracted and used as seeds. Ground-truth domain labels were derived directly from the researcher-assigned themes, preserving independence from LLM outputs:

- Vent-Schmidt J, Attara GP, Lisko D, Steiner TS. Patient Experiences with Clostridioides difficile Infection: Results of a Canada-Wide Survey. Patient Prefer Adherence. 2020;14:33-43. PMID 32021115. CDI-specific; themes included diagnostic frustration, fear of recurrence, and social isolation.
- Barnes EL, Boynton MH, DeWalt DA, Brenner E, Herfarth HH, Kappelman MD. Understanding the Lived Experience After Colectomy and Ileal Pouch-Anal Anastomosis for Ulcerative Colitis: A Qualitative Study. Crohns Colitis 360. 2025;7(1):otaf007. PMID 39936138. Related gastrointestinal/colitis source; themes included treatment fatigue, surgical decision-making, and caregiver role.
- Patel RK, Teja R, Hermann K, Franz R, Wong K, Kao D. Engaging Patient and Caregiver Partners in Codeveloping a Patient Educational Video for Improving Clostridioides difficile Infection Education: Participatory Co-Design Study. JMIR Form Res. 2026;10(1):e81643. PMID 41780918. Recurrent CDI/FMT patient-education co-design; themes included patient agency, information barriers, and FMT decision-making.

#### S4.3 Corpus Generation

Claude Sonnet 4.6 was used at a temperature of 0.8 to generate 60 synthetic testimonials (220-290 words each) seeded from the extracted quotes. Each item was assigned a blueprint specifying the cohort (rCDI or fulminant), target domains to include (based on the source paper's themes), a primary arc, and a word-count range. The model was instructed to write a plausible first-person or caregiver testimonial that authentically reflected the specified themes. The generator was not told which domain labels to activate: it received thematic descriptions, and the model's realization determined the final presence/absence of each domain.

The 60-item corpus was split: items 1, 4, 7, ... (every 3rd, starting at index 0) were assigned to the held-out validation set (n=20); the remaining 40 items formed the tuning set.

### S5. Three-Iteration Prompt Tuning

#### S5.1 Iterations Tested

Three prompt iterations were evaluated on the 40-item tuning set:

- Iteration 0 (Baseline): Standard system prompt with domain definitions, arc typology, and JSON output format. No threshold calibration.
- Iteration 1 (+Calibration): Baseline plus the D5/D6/D7 calibration addendum reproduced in S1.2. Addressed the primary source of inter-model disagreement identified in baseline testing.
- Iteration 2 (+Model-specific): Iteration 1 plus model-specific reminder addenda. For Claude: additional reminder that D7=0 when the patient passively follows the clinician's advice. For DeepSeek: additional reminder that D6 requires explicit friction, not routine care.

#### S5.2 Objective Metric

The objective metric was macro-F1 averaged over the 8 standard domains (D1-D8), computed as the unweighted mean of per-domain F1 scores against the researcher-assigned ground truth. Fulminant-specific domains (F1-F4) were excluded from the tuning objective because the synthetic benchmark was lightly weighted toward fulminant items and ground-truth labels for F-domains had lower confidence in the literature sources.

#### S5.3 Tuning Results

**Table SM3. Three-Iteration Prompt Tuning Results**

| Iteration | Claude macro-F1 (tuning set, n=40) | DeepSeek macro-F1 (tuning set, n=40) | Notes |
|---|---|---|---|
| 0 -- Baseline | 0.717 | 0.847 | Pre-calibration baseline |
| 1 -- +Calibration | 0.792 | 0.855 | D5/D6/D7 threshold addendum added |
| 2 -- +Model-specific | 0.790 | 0.851 | Marginal or negative vs. iter 1; iter 1 selected |

*Macro-F1 = unweighted mean of per-domain F1 across D1-D8. Iteration 1 was selected as optimal for both models (parsimony principle: identical tuning set for both models). Iteration 2 showed marginal or slightly negative gain, suggesting the D5/D6/D7 addendum captured the primary improvement signal.*

### S6. Held-Out Validation and GO/NO-GO Decision

#### S6.1 Protocol

The 20-item held-out validation set was used to assess the generalization of the tuned iteration-1 prompt. Items were coded by both models using the iteration-1 prompt. Macro-F1 on the validation set was compared to: (a) the baseline tuning F1 (iteration 0 on the tuning set) to compute improvement; and (b) the best-iteration tuning F1 (iteration 1 on the tuning set) to detect overfitting.

Pre-specified decision thresholds:

- GO: validation F1 improvement >=0.05 vs. baseline AND validation F1 >= (tuning F1 minus 0.05)
- NO-GO: either threshold not met

#### S6.2 Validation Results

**Table SM4. Held-Out Validation GO/NO-GO Decision**

| Model | Baseline tuning F1 | Iter-1 tuning F1 | Validation F1 (n=20) | Decision |
|---|---|---|---|---|
| Claude Sonnet 4.6 | 0.717 | 0.792 | 0.815 | GO |
| DeepSeek Chat | 0.847 | 0.855 | 0.903 | GO |

*Improvement vs. baseline: Claude +0.098 (threshold >=0.05 met); DeepSeek +0.056 (threshold met). Both models showed positive generalization (validation F1 exceeded tuning F1), ruling out over-fit. GO decision confirmed for both models.*

#### S6.3 Per-Domain Validation F1

**Table SM5. Per-Domain Validation F1 (n=20)**

| Domain | Domain Name | Claude F1 | DeepSeek F1 | Note |
|---|---|---|---|---|
| D1 | Diagnostic journey | 1.000 | 0.857 | |
| D2 | Treatment trajectory | 0.600 | 1.000 | Small n positive; both adequate |
| D3 | Physical symptom burden | 0.923 | 1.000 | |
| D4 | Psychological/ emotional burden | 0.737 | 0.824 | |
| D5 | Social/functional disruption | 0.857 | 0.857 | Calibration effective |

| | | | | |
|---|---|---|---|---|
| D6 | Healthcare system experience | 0.933 | 0.933 | Calibration effective |
| D7 | Information-seeking/agency | 0.667 | 0.750 | Lowest domain; acceptable |
| D8 | Recovery/resilience | 0.800 | 1.000 | |
| F1 | Sepsis/organ failure | 1.000 | 1.000 | Fulminant subset only (~n=8) |
| F2 | ICU/life support | 1.000 | 1.000 | |
| F3 | Major surgery | 1.000 | 1.000 | |
| F4 | Death/near-death | 1.000 | 1.000 | |
| | **Macro-F1 (standard domains, D1-D8)** | **0.815** | **0.903** | |

*F1 computed per-domain against researcher-assigned ground truth (n=20 held-out items). Fulminant-domain n is approximate (varies by blueprint). D7 had the lowest validation F1 for both models; this is consistent with its inherently contextual nature and the active-agency threshold.*

### S7. Inter-rater Reliability Protocol

#### S7.1 Subsample Selection

A stratified random subsample of 16 testimonials (IRR-001 through IRR-016; 14 rCDI + 2 fulminant; 376-656 words; random seed=42) was selected for human coding. Stratification ensured: (a) proportional representation of word-count quartiles; (b) inclusion of at least 2 fulminant items; (c) exclusion of items shorter than 350 words (insufficient content for reliable domain assessment).

#### S7.2 Human Coders

Two independent human coders, Coder 1 and Coder 2, both senior internal medicine residents, applied the full codebook independently and blind to each other's ratings and to LLM outputs during primary coding. Each coder received a standardized coding packet including: full testimonial text (one item per page, formatted for paper use); paper coding forms with domain checkboxes and arc selection; a CDI domain primer and methods guide; the codebook (Table SM1); and a practice set with answer key. Coders contributed to the design of IRR analyses and will review manuscript drafts.

#### S7.3 Calibration Session and Post-Calibration Recoding

Following primary coding, a calibration session was convened to discuss disagreements on a subset of items. Seven items were identified for post-calibration recoding (IRR-001, IRR-003, IRR-009, IRR-010, IRR-011, IRR-013, IRR-015). Each coder independently submitted revised codings for these 7 items after calibration.

#### S7.4 PI Adjudication

Remaining disagreements after post-calibration recoding were adjudicated by the PI. The PI reviewed the full testimonial text and all coder responses before assigning a final reference standard. The PI adjudicated 28 field-level disagreements across 7 of the 16 IRR items, each row representing one dimension decision on one testimonial. This final PI adjudication constitutes the reference standard for the 16-item IRR subsample.

#### S7.5 Agreement Metrics

**PABAK (primary metric):** [3]

**Gwet's AC1:** [4]

**Cohen's kappa:** [5]

**Landis-Koch interpretation:** [6]

Because the inter-rater subsample is small (n=16), agreement is reported as point estimates without confidence intervals.

#### S7.6 Three-Way Results and LLM-LLM Arm

Three-way concordance (Coder 1, Coder 2, Claude Sonnet 4.6) across the 16-item IRR subsample is reported in Supplementary Table S3. Human-human pairwise agreement before and after calibration and LLM-human pairwise comparisons are reported in Supplementary Table S3.

Reproducibility asymmetry, the pattern in which human coders agreed more with each other than either agreed with the LLM on certain dimensions, is detailed in Supplementary Table S4. Cluster A dimensions (with higher human-human than LLM-LLM agreement) reflect dimensions that are more dependent on clinical contextual judgment; Cluster B dimensions (with higher LLM-LLM agreement) reflect dimensions in which structured codebook thresholds are more determinative.

LLM-LLM concordance between Claude Sonnet 4.6 and DeepSeek V4 Pro was computed on the full analytic corpus (n=189). Results are incorporated into Supplementary Table S3.

Post-calibration improvement: human-human PABAK improved from 0.780 to 0.912 on SF dimensions following the calibration session and 7-item recoding. Binary domain (D-dimension) results and narrative arc diverged further post-calibration in some pairs; PI adjudication of the 28 remaining field-level disagreements established the final reference standard.

**Supplemental Table S1. Full coding frequencies (%) by CDI cohort.**

*Cohort columns show per-model coding as Claude/DeepSeek (%); the Total column is the pooled dual-model mean. F1–F4 flags were applied only to fulminant cohorts (rfCDI, fCDI); non-fulminant cohorts are not applicable (—). The total for F-flags is weighted by rfCDI + fCDI (n=67).*

| Coding dimension | rfCDI (n=49) | fCDI (n=18) | rCDI (n=84) | non-rfCDI (n=38) | Total (n=189) |
|---|---|---|---|---|---|
| ***Qualitative coding domains (D1–D8)*** | | | | | |
| D1 — Diagnostic journey | 88/86 | 78/50 | 92/76 | 100/76 | **83.7%** |
| D2 — Treatment trajectory | 100/100 | 72/67 | 100/100 | 97/97 | **96.6%** |
| D3 — Physical symptom burden | 100/100 | 94/72 | 100/93 | 100/97 | **96.6%** |
| D4 — Psychological/emotional burden | 98/98 | 94/100 | 92/92 | 74/66 | **89.5%** |
| D5 — Social/functional disruption | 78/78 | 44/39 | 60/58 | 45/58 | **60.6%** |
| D6 — Healthcare system experience | 98/98 | 78/89 | 90/89 | 74/82 | **88.9%** |
| D7 — Info-seeking / patient agency | 86/82 | 61/56 | 80/76 | 66/66 | **75.2%** |
| D8 — Recovery, resilience and outlook | 78/71 | 39/39 | 69/62 | 68/55 | **64.6%** |
| ***Fulminant severity flags (F1–F4; rfCDI and fCDI only)*** | | | | | |
| F1 — Sepsis / organ failure | 57/57 | 67/67 | — | — | **59.7%** |
| F2 — ICU / ventilation | 14/14 | 39/33 | — | — | **20.2%** |
| F3 — Emergency surgery | 16/20 | 39/39 | — | — | **23.9%** |
| F4 — Death / near-death | 33/39 | 83/78 | — | — | **47.8%** |
| ***Semantic fields (SF01–SF13)*** | | | | | |
| SF01 — Antibiotic trigger | 67/74 | 56/61 | 76/75 | 79/79 | **73.3%** |
| SF02 — Diagnostic failure | 51/61 | 33/39 | 44/51 | 40/40 | **47.1%** |
| SF03 — Bacteriotherapy | 61/59 | 6/6 | 46/44 | 8/5 | **37.6%** |
| SF04 — Financial / insurance burden | 31/31 | 11/11 | 32/36 | 13/13 | **26.7%** |
| SF05 — Physical severity | 100/100 | 94/100 | 77/87 | 63/76 | **85.8%** |
| SF06 — Psychological / mental health | 63/82 | 33/50 | 55/69 | 26/37 | **56.6%** |
| SF07 — Social / occupational disruption | 57/55 | 17/28 | 39/43 | 24/34 | **40.8%** |
| SF08 — Surgical / critical intervention | 26/22 | 44/56 | 4/6 | 3/5 | **14.0%** |
| SF09 — Healthcare system failure | 71/69 | 56/56 | 56/64 | 50/53 | **60.6%** |
| SF10 — Patient agency / research | 69/59 | 28/28 | 67/62 | 53/50 | **58.2%** |
| SF11 — Caregiver / bereavement | 12/12 | 67/67 | 1/4 | 0/0 | **10.6%** |
| SF12 — Long-term sequelae | 65/71 | 28/28 | 43/42 | 40/42 | **47.4%** |
| SF13 — Diet change / probiotic | 33/33 | 17/17 | 42/43 | 45/47 | **38.1%** |

*Abbreviations: rfCDI, recurrent fulminant CDI (n=49); fCDI, fulminant non-recurrent CDI (n=18); rCDI, recurrent non-fulminant CDI (n=84); non-rfCDI, non-recurrent non-fulminant CDI (n=38). D1–D8, qualitative coding domains; F1–F4, fulminant severity flags; SF01–SF13, semantic fields. —, not applicable.*

**Supplemental Table S2. Pre- vs post-calibration concordance (Coder 1 vs Coder 2, 7-item calibration subset).**

*Agreement between the two human coders on the 7-item calibration subset, before and after a consensus discussion. F-flag rows (F1–F4) include only the three fulminant-CDI items (n=3). Δ PABAK = post − pre.*

| Coding dimension | n | Pre PABAK | Post PABAK | Δ PABAK | Pre AC1 | Post AC1 |
|---|---|---|---|---|---|---|
| ***Binary coding dimensions (D1–D8, F1–F4)*** | | | | | | |
| D1 — Diagnostic journey | 7 | 1.000 | 0.429 | -0.571 | 1.000 | 0.517 |
| D2 — Treatment trajectory | 7 | 0.714 | 0.143 | -0.571 | 0.835 | 0.354 |
| D3 — Physical symptom burden | 7 | 0.714 | 0.714 | +0.000 | 0.835 | 0.835 |
| D4 — Psychological/emotional burden | 7 | 0.714 | 1.000 | +0.286 | 0.835 | 1.000 |
| D5 — Social/functional disruption | 7 | 0.714 | 0.429 | -0.285 | 0.835 | 0.622 |
| D6 — Healthcare system experience | 7 | 0.429 | 0.429 | +0.000 | 0.622 | 0.622 |
| D7 — Info-seeking / patient agency | 7 | 1.000 | 1.000 | +0.000 | 1.000 | 1.000 |
| D8 — Recovery, resilience and outlook | 7 | 0.429 | 0.429 | +0.000 | 0.517 | 0.440 |
| F1 — Sepsis / organ failure | 3 | -0.333 | -0.333 | +0.000 | -0.200 | -0.200 |
| F2 — ICU / ventilation | 3 | 1.000 | 1.000 | +0.000 | 1.000 | 1.000 |
| F3 — Emergency surgery | 3 | 1.000 | 0.333 | -0.667 | 1.000 | 0.333 |
| F4 — Death / near-death | 3 | 1.000 | 1.000 | +0.000 | 1.000 | 1.000 |
| **Mean** | | **0.698** | **0.548** | **-0.150** | **0.773** | **0.627** |
| ***Semantic fields (SF01–SF13)*** | | | | | | |
| SF01 — Antibiotic trigger | 7 | 0.714 | 1.000 | +0.286 | 0.736 | 1.000 |
| SF02 — Diagnostic failure | 7 | 0.429 | 0.714 | +0.285 | 0.517 | 0.736 |
| SF03 — Bacteriotherapy | 7 | 1.000 | 1.000 | +0.000 | 1.000 | 1.000 |
| SF04 — Financial / insurance burden | 7 | 1.000 | 1.000 | +0.000 | 1.000 | 1.000 |
| SF05 — Physical severity | 7 | 1.000 | 1.000 | +0.000 | 1.000 | 1.000 |
| SF06 — Psychological / mental health | 7 | 1.000 | 1.000 | +0.000 | 1.000 | 1.000 |
| SF07 — Social / occupational disruption | 7 | 1.000 | 0.429 | -0.571 | 1.000 | 0.622 |
| SF08 — Surgical / critical intervention | 7 | 1.000 | 1.000 | +0.000 | 1.000 | 1.000 |
| SF09 — Healthcare system failure | 7 | 0.429 | 1.000 | +0.571 | 0.517 | 1.000 |
| SF10 — Patient agency / research | 7 | 0.714 | 1.000 | +0.286 | 0.785 | 1.000 |
| SF11 — Caregiver / bereavement | 7 | 1.000 | 1.000 | +0.000 | 1.000 | 1.000 |
| SF12 — Long-term sequelae | 7 | 0.429 | 0.714 | +0.285 | 0.440 | 0.785 |
| SF13 — Diet change / probiotic | 7 | 0.429 | 1.000 | +0.571 | 0.440 | 1.000 |
| **Mean** | | **0.780** | **0.912** | **+0.132** | **0.803** | **0.934** |

| PABAK > 0.80 — Almost perfect | PABAK 0.61-0.80 — Substantial | PABAK 0.41-0.60 — Moderate | PABAK 0.21-0.40 — Fair | PABAK 0.00-0.20 — Slight / Poor |
|---|---|---|---|---|

*Abbreviations: PABAK, prevalence-adjusted bias-adjusted kappa (Byrt 1993); AC1, Gwet's first-order agreement coefficient; Δ, change (post − pre). F-flags (F1–F4) include only fulminant-CDI items (n=3).*

**Supplemental Table S3. Inter-rater reliability concordance by coding dimension. LLM x LLM: full analytic corpus (Claude Sonnet 4.6 vs DeepSeek V4; D1-D8 n=189, F1-F4 n=67 fulminant, SF n=189). Human comparisons: IRR subsample (n=16, post-PI-adjudication codings).**

**Panel A: Binary coding dimensions (D1-D8: LLM-LLM n=189, human n=16; F1-F4: LLM-LLM n=67, human n=6 fulminant items only)**

| | | ***LLM x LLM (Claude x DeepSeek)*** | | | ***Human x Human (Coder 1 x Coder 2)*** | | | ***Human x LLM (Coder 1 x LLM)*** | | | ***Human x LLM (Coder 2 x LLM)*** | | |
|---|---|---|---|---|---|---|---|---|---|---|---|---|---|
| **Dimension** | **n** | **PABAK** | **κ** | **AC1** | **PABAK** | **κ** | **AC1** | **PABAK** | **κ** | **AC1** | **PABAK** | **κ** | **AC1** |
| D1 — Diagnostic journey | 16 | **0.66** | 0.41 | 0.77 | **0.88** | 0.64 | 0.93 | **0.50** | —† | 0.68 | **0.62** | —† | 0.77 |
| D2 — Treatment trajectory | 16 | **0.99** | 0.92 | 0.99 | **0.88** | —† | 0.93 | **0.75** | —† | 0.86 | **0.88** | —† | 0.93 |
| D3 — Physical symptom burden | 16 | **0.88** | —† | 0.94 | **0.88** | —† | 0.93 | **0.88** | —† | 0.93 | **1.00** | 1.00 | 1.00 |
| D4 — Psychological/emotional burden | 16 | **0.89** | 0.72 | 0.94 | **0.62** | 0.48 | 0.71 | **0.75** | —† | 0.86 | **0.38** | —† | 0.58 |
| D5 — Social/functional disruption | 16 | **0.76** | 0.74 | 0.77 | **0.38** | 0.31 | 0.47 | **0.25** | 0.19 | 0.34 | **0.62** | —† | 0.74 |
| D6 — Healthcare system experience | 16 | **0.85** | 0.62 | 0.91 | **0.75** | —† | 0.86 | **0.62** | —† | 0.74 | **0.62** | —† | 0.77 |
| D7 — Info-seeking and patient agency | 16 | **0.87** | 0.83 | 0.90 | **0.75** | 0.59 | 0.82 | **0.50** | —† | 0.68 | **0.75** | —† | 0.84 |
| D8 — Recovery, resilience and outlook | 16 | **0.83** | 0.82 | 0.84 | **0.62** | 0.61 | 0.64 | **0.62** | 0.64 | 0.63 | **0.50** | 0.53 | 0.50 |
| F1 — Sepsis / organ failure | 6 | **0.82** | 0.81 | 0.83 | **0.33** | —† | 0.54 | **0.67** | 0.57 | 0.73 | **0.67** | —† | 0.80 |
| F2 — ICU / ventilation | 6 | **0.97** | 0.95 | 0.98 | **1.00** | 1.00 | 1.00 | **1.00** | 1.00 | 1.00 | **1.00** | 1.00 | 1.00 |
| F3 — Emergency surgery | 6 | **0.94** | 0.92 | 0.95 | **1.00** | 1.00 | 1.00 | **1.00** | 1.00 | 1.00 | **1.00** | 1.00 | 1.00 |
| F4 — Death / near-death | 6 | **0.58** | 0.58 | 0.58 | **1.00** | 1.00 | 1.00 | **1.00** | 1.00 | 1.00 | **1.00** | 1.00 | 1.00 |
| **Mean** | | **0.838** | — | 0.866 | **0.757** | — | 0.820 | **0.712** | — | 0.788 | **0.753** | — | 0.829 |

**Panel B: Emerging semantic fields (SF01-SF13; LLM-LLM n=189, human n=16)**

| | | ***LLM x LLM (Claude x DeepSeek)*** | | | ***Human x Human (Coder 1 x Coder 2)*** | | | ***Human x LLM (Coder 1 x LLM)*** | | | ***Human x LLM (Coder 2 x LLM)*** | | |
|---|---|---|---|---|---|---|---|---|---|---|---|---|---|
| **Dimension** | **n** | **PABAK** | **κ** | **AC1** | **PABAK** | **κ** | **AC1** | **PABAK** | **κ** | **AC1** | **PABAK** | **κ** | **AC1** |
| SF01 — Antibiotic Trigger | 16 | **0.93** | 0.91 | 0.94 | **0.75** | 0.74 | 0.77 | **0.75** | 0.74 | 0.77 | **1.00** | 1.00 | 1.00 |
| SF02 — Diagnostic Failure | 16 | **0.83** | 0.83 | 0.83 | **0.50** | 0.51 | 0.50 | **0.25** | 0.28 | 0.26 | **0.75** | 0.74 | 0.77 |
| SF03 — Bacteriotherapy | 16 | **0.94** | 0.93 | 0.94 | **1.00** | 1.00 | 1.00 | **1.00** | 1.00 | 1.00 | **1.00** | 1.00 | 1.00 |

| | | | | | | | | | | | | | |
|---|---|---|---|---|---|---|---|---|---|---|---|---|---|
| SF04 — Financial / Insurance Burden | 16 | **0.91** | 0.88 | 0.92 | **1.00** | 1.00 | 1.00 | **1.00** | 1.00 | 1.00 | **1.00** | 1.00 | 1.00 |
| SF05 — Physical Severity | 16 | **0.85** | 0.70 | 0.90 | **0.88** | 0.64 | 0.93 | **1.00** | 1.00 | 1.00 | **0.88** | 0.64 | 0.93 |
| SF06 — Psychological / Mental Health | 16 | **0.68** | 0.68 | 0.69 | **0.88** | 0.85 | 0.90 | **0.12** | 0.26 | 0.13 | **0.25** | 0.34 | 0.25 |
| SF07 — Social / Occupational Disruption | 16 | **0.79** | 0.78 | 0.80 | **0.75** | 0.71 | 0.78 | **0.25** | 0.33 | 0.26 | **0.00** | 0.20 | 0.00 |
| SF08 — Surgical / Critical Intervention | 16 | **0.93** | 0.85 | 0.95 | **1.00** | 1.00 | 1.00 | **0.75** | 0.67 | 0.80 | **0.75** | 0.67 | 0.80 |
| SF09 — Healthcare System Failure | 16 | **0.82** | 0.81 | 0.83 | **0.50** | 0.50 | 0.51 | **0.25** | 0.25 | 0.26 | **0.75** | 0.75 | 0.75 |
| SF10 — Patient Agency / Research | 16 | **0.83** | 0.83 | 0.83 | **0.75** | 0.59 | 0.82 | **0.25** | 0.30 | 0.29 | **0.25** | 0.30 | 0.29 |
| SF11 — Caregiver / Bereavement | 16 | **0.98** | 0.94 | 0.99 | **1.00** | 1.00 | 1.00 | **0.88** | 0.86 | 0.89 | **0.88** | 0.86 | 0.89 |
| SF12 — Long-term Sequelae | 16 | **0.86** | 0.86 | 0.86 | **0.50** | 0.46 | 0.56 | **0.50** | 0.46 | 0.56 | **1.00** | 1.00 | 1.00 |
| SF13 — Diet Change / Probiotic | 16 | **0.94** | 0.93 | 0.94 | **0.75** | 0.71 | 0.78 | **0.75** | 0.71 | 0.78 | **1.00** | 1.00 | 1.00 |
| **Mean** | | **0.867** | — | 0.879 | **0.788** | — | 0.811 | **0.596** | — | 0.616 | **0.731** | — | 0.744 |

PABAK > 0.80 — Almost perfect    PABAK 0.61-0.80 — Substantial    PABAK 0.41-0.60 — Moderate
PABAK 0.21-0.40 — Fair    PABAK 0.00-0.20 — Slight / Poor

**PABAK:** Prevalence-Adjusted Bias-Adjusted Kappa (Byrt 1993). Primary metric: robust to prevalence paradox. Landis & Koch thresholds applied.

**AC1:** Gwet's AC1 (Gwet 2008). Alternative chance-correction robust to prevalence paradox; reported for convergent validity.

**κ:** Cohen's kappa. Where |kappa - PABAK| > 0.30 (prevalence paradox), kappa is shown as —† and should not be interpreted; PABAK and AC1 are preferred.

**H x H:** Human x Human (Coder 1 x Coder 2): primary inter-rater reliability estimate.

**LLM x LLM:** Claude Sonnet 4.6 vs DeepSeek V4 (DeepSeek-Chat); full analytic corpus (D1-D8 n=189, F1-F4 n=67 fulminant only, SF n=189). Both models run at temperature 0; codings are independent (no cross-model feedback).

**H x LLM:** Human x LLM: alignment of each human coder independently with the LLM reference coder (Claude Sonnet 4.6).

**Arc / dom:** Arc and dominant-dimension agreement are exploratory; percent agreement: Coder 1 x Coder 2 68.8%, Coder 1 x LLM 37.5%, Coder 2 x LLM 43.8%.

**Supplemental Table S4. Dimensions of asymmetric inter-rater reproducibility between human and model coders. Values are PABAK unless marked; model vs model is raw Claude Sonnet 4.6 against raw DeepSeek V4, human vs human is coder 1 vs. 2.**

##### Cluster A. Human raters are more reproducible than the models

*On these dimensions, the two clinicians agreed with each other more than the two models did, which may reflect clinical training applied to content that requires interpretation beyond the explicit text.*

| Dimension | Model vs model (n=16) | Model vs model (n=189) | Human vs human (n=16) | Speculated mechanism |
|---|---|---|---|---|
| SF07 Social / Occupational Disruption | 0.25 | 0.79 | **0.75** | Both coders reliably read indirect cues, such as an inability to work or being housebound, as disruptions. The models recovered this implicit signal inconsistently, agreeing with each other only weakly on the hard items. |
| SF06 Psychological / Mental Health | 0.50 | 0.68 | **0.88** | Clinicians appear to infer psychological burden from tone and described disruption and agree on it, while the models required explicit mental-health language and varied when it was absent. |
| D1 Diagnostic journey | 0.63 | 0.66 | **0.88** | Both coders are senior internal medicine residents, for whom interpreting diagnostic and therapeutic journeys, including their complexities and pitfalls, is routine clinical work. This shared expertise may support a consistent reading of the diagnostic-journey narrative that the models reproduced only moderately at either sample size. |
| SF10 Patient Agency / Research | 0.63 | 0.83 | **0.75** | A weaker form of the same pattern: the coders recognized self-advocacy in the described actions somewhat more consistently than the models did. |
| Narrative arc † | 50.0% | — | **68.8%** | The coders converged on a single arc more often than the models did, consistent with a shared reading of the trajectory. Exploratory. |

##### Cluster B. Models are more reproducible than human raters

*On these dimensions, the two models, run deterministically at temperature 0, reproduced one another more consistently than the clinicians, who varied in where they set a threshold or which single frame they selected.*

| Dimension | Model vs model (n=16) | Model vs model (n=189) | Human vs human (n=16) | Speculated mechanism |
|---|---|---|---|---|
| SF12 Long-term Sequelae | **1.00** | 0.86 | 0.50 | Humans differed on what counts as long-term, ranging from lingering symptoms to explicit chronicity. The models applied a consistent threshold and agreed completely on the edge items. |
| SF02 Diagnostic Failure | **0.75** | 0.83 | 0.50 | Humans split on the threshold for diagnostic failure, with one coder counting any reported delay. The models applied it consistently. |
| SF09 Healthcare System Failure | **0.75** | 0.82 | 0.50 | Humans differed on whether system failure requires a structural failure or includes an individual clinician error; the models were consistent. |
| D4 Psychological / emotional burden | **0.88** | 0.89 | 0.63 | The broad emotional-burden domain was coded reproducibly by the models but variably by humans. The narrow field SF06 shows the opposite, so the effect appears domain-specific. |
| D8 Recovery, resilience and outlook | **0.88** | 0.83 | 0.63 | A whole-narrative judgment of recovery drifted between human readers but was reproduced by the deterministic models. |
| SF13 Diet change / probiotic | **1.00** | 0.94 | 0.75 | The models were fully reproducible; the coders were substantial but not perfect, differing on borderline dietary and probiotic mentions. |
| Dominant dimension † | **62.5%** | — | 37.5% | Selecting one dominant frame was reproduced by the models far more often than by the coders, and is the basis of Figure 4. The coders agreed with the model more often than with each other. |

**Note.** D5 Social/functional disruption is not listed in either cluster. Agreement was low for both rater types on the edge items (models: 0.50; humans: 0.38), and the larger full-corpus model value (0.76) does not hold for the harder cases. D5 is thus a target for instrument refinement.

(1) PABAK, prevalence-adjusted bias-adjusted kappa (Byrt 1993).

(2) The n=16 set is the inter-rater reliability subsample selected to represent coding difficulty; n=189 is the full analytic corpus. Edge items are harder, so full-corpus model agreement runs higher. D5 illustrates this difference (0.76 corpus, 0.50 edge).

(3) † Arc and dominant dimension are multi-class and reported as percent exact agreement, not PABAK.

(4) The fulminant flags F1 to F4 rest on ten or fewer items at n=16 and are not interpreted here.

**Supplemental Table S5. Representative patient testimonial excerpts by semantic field (SF01–SF13).**

*Excerpts are drawn from PI-reviewed testimonials; names, clinician identifiers, and institution names have been removed or generalized. Cohort abbreviations: rfCDI = recurrent fulminant CDI; fCDI = fulminant non-recurrent CDI; rCDI = recurrent non-fulminant CDI.*

**SF01 — Antibiotic Trigger**

| Excerpt | Cohort |
|---|---|
| *"I was prescribed several rounds of antibiotics by health care professionals for what they diagnosed as SIBO (whether I actually had SIBO or not, I still don't know). Due to those antibiotics, I was dealt one of the worst cases of C. diff doctors in the Boston area had ever seen in someone my age. Six months and four rounds of the strongest antibiotics around to try and stop the infection to no avail."* | rfCDI |
| *"After a visit to my Primary Care Provider (PCP) for a possible sinus infection, I was prescribed antibiotics and referred to my dentist for a possible tooth problem. Two weeks later, I saw my dentist who did some dental work and prescribed clindamycin. No sooner than I completed the antibiotics, I woke up in the middle of a Friday night with chills, fever, vomiting, and uncontrollable diarrhea."* | rCDI |

**SF02 — Diagnostic Failure**

| Excerpt | Cohort |
|---|---|
| *"After several hours, I finally saw a doctor, who ordered an ultrasound to […] rule out that what I was experiencing was not due to an ectopic pregnancy or a miscarriage. They did not run any other tests, despite my asking, as they were just too busy with their one other patient in the ER that night. I continued to decline physically, and after several rounds of fluids [..], I was sent home with a general diagnosis of a stomach bug, saying it will clear up on it's own."* | fCDI |
| *"It took until June 30, 2025 to be diagnosed because the gastroenterologist I saw twice insisted there was absolutely nothing wrong with me, refusing to run any tests because it would be "a waste of time." My present gastroenterologist was the one to diagnose me and is now treating me."* | rCDI |

**SF03 — Bacteriotherapy**

| Excerpt | Cohort |
|---|---|
| *"They researched my history and sent a full review of what they felt was best for me, and they recommended a fecal microbiota transplant (FMT). So on my fifth stay in the hospital, the doctors read the recommendation and put me in contact with the only gastrointestinal doctor here in Las Vegas that performs FMTs."* | rfCDI |

**SF04 — Financial / Insurance Burden**

| Excerpt | Cohort |
|---|---|
| *"I have no savings, no career and am currently fighting for approval of permanent disability. I've lost my health insurance and am now on Medicaid, living in my childhood home that my family owns and lets me live in. This disease is so brutal that without my family and Boyfriend to save me, C. diff would have also made me homeless."* | rfCDI |
| *"I was able to get insurance through the Affordable Care Act beginning on January 1, 2014. However, this has its restrictions; if I earn more than $150 in any single month, it compromises my eligibility for the insurance. I have spent all of my personal resources (money that I had saved for law school and my daughter's wedding), and I now face uncertain financial times because I was also recently denied disability."* | rCDI |

**SF05 — Physical Severity**

| Excerpt | Cohort |
|---|---|

| | |
|---|---|
| *"Upon leaving, I was prescribed Clindamycin (a strong antibiotic) in the event I develop an infection. Mild pain and diarrhea soon began to interrupt my daily routine, which quickly escalated into debilitating pain, severe dehydration, and uncontrollable diarrhea. I was hospitalized and diagnosed with C. diff, which I had never heard of before."* | rfCDI |
| *"I began having diarrhea that evening, and it progressed throughout the night and the next day, Sunday. By Sunday night, the 28th, I was on the floor of our living room with debilitating stomach pain after a recent trip to the bathroom. I was taken to the Emergency Room […], I had severe pain and diarrhea at that point, and was also extremely dehydrated."* | fCDI |

**SF06 — Psychological / Mental Health**

| Excerpt | Cohort |
|---|---|
| *"But what most people don't talk about is the emotional side of C. diff. The anxiety, panic over every gurgle or bathroom trip, and the isolation that comes from being scared to leave home have been incredibly hard to cope with. I'm currently in therapy because I've realized I've developed a fear of going out, something I never struggled with before."* | fCDI |
| *"This is one of the reasons I left my job and decided to work from home. You are never fully warned about the long-term damage that antibiotics can cause to the gut flora or the traumatic experience of C. difficile. What they don't tell you about is the emotional trauma (it's true, we all stress we are going to have a relapse), the financial loss or the stress of managing a job whilst being severely ill, and the long term lasting effects that you have to deal with after."* | rCDI |

**SF07 — Social / Occupational Disruption**

| Excerpt | Cohort |
|---|---|
| *"It also had negative effects on my relationships with my family and friends. I planned to go back to work after three months but had to cancel all of my clients because I was suffering with a recurrent infection. When I could finally return after six months I had lost a lot of my client base that I had spent so long building."* | rfCDI |
| *"I couldn't go to class or work and I barely left my bed — only to use the bathroom. I was enrolled full-time at a University and working two jobs and I could barely push through due to the pain I was experiencing. I felt like my friends, coworkers, professors, and family members could not understand what was going on, and when I explained I just felt embarrassed and humiliated by my awful side effects."* | rCDI |

**SF08 — Surgical / Critical Intervention**

| Excerpt | Cohort |
|---|---|
| *"I spent three weeks in a drug-induced coma while on a ventilator. I had four surgeries while I was in the ICU; the first was a removal of my entire colon, in which I was given an ileostomy. The second surgery was to open up my abdomen in an attempt to reduce the pressure on my organs, which were failing due to the effects of severe sepsis and abdominal compartment syndrome."* | rfCDI |

**SF09 — Healthcare System Failure**

| Excerpt | Cohort |
|---|---|
| *"I knew that I must have been dehydrated, and at some point I was no longer able to urinate, despite having ingested plenty of liquids. I went to the emergency room, and was finally seen after being ignored in the waiting room for 45 minutes. I was admitted by the triage nurse "stat with emergent code blue." She couldn't even get a blood pressure reading from me."* | rfCDI |
| *"My battle with C. diff started when I cut my ankle and was prescribed multiple antibiotics to prevent infection. About a week after finishing my last antibiotic, I began experiencing symptoms of C. diff. I visited a gastroenterologist, and while waiting for test results, she advised me to take Immodium, which I later discovered was the most detrimental course of action."* | rCDI |

**SF10 — Patient Agency / Research**

| Excerpt | Cohort |
|---|---|
| *"When my journey began, I was completely unaware of C. diff. Now, I have an understanding of the disease, became my own advocate and through my own research, learned effective strategies to combat it. While my story may not be as extraordinary as others I've read about, I firmly believe that my proactive approach and self-advocacy played a pivotal role in achieving what has so far been a favorable outcome."* | rCDI |

**SF11 — Caregiver / Bereavement**

| Excerpt | Cohort |
|---|---|
| *"The very woman I made sure I spent time with and planned to spend more time with after I retired was sick, but I didn't know how serious it was until I spent days and hours with her in the hospital alternating shifts with my father from 9am – 9am. My father loved my mother so much he wouldn't leave her side or take a break. My mother would have to force him to go eat because she worried about him."* | rfCDI |
| *"I am writing this story for my Mom […] My mom was 64 years young when she died from C. diff colitis on August 5, 2012. I was 27 years old at the time. My mom had long suffered from chronic obstructive pulmonary disease (COPD) and bouts of pneumonia since my father passed away of cancer in 2009."* | fCDI |
| *"She had an Ileostomy done in March 2009. It saved her life, but she was never the same mentally or physically. For seven years, she suffered with multiple illnesses and, ultimately, passed away on November 16, 2016."* | rCDI |

**SF12 — Long-term Sequelae**

| Excerpt | Cohort |
|---|---|
| *"Now, 2024, it's been 5 years since my 2nd transplant. 5 years later I am no longer hoping or praying for healing. I know that isn't coming now. Now, I just pray that I will be approved for permanent disability so I am no longer a financial burden to my family and Boyfriend."* | rfCDI |
| *"It was the worst illness I've ever had and to this day, 17 years later, I'm still deathly afraid of taking any medication or being around sick people. I learned my lesson but still, it's scary how so many people don't wash their hands and they can transmit this deathly disease to others. I have to take a probiotic (Culturelle) every day or else my stomach is upset."* | rCDI |

**SF13 — Diet Change / Probiotic**

| Excerpt | Cohort |
|---|---|
| *"My sister pleaded to have her discharged with an adequate supply of vancomycin to allow for a slow titration down from treatment. Although the rehab facility had been administering lactobacillus twice daily, I learned only Lactobacillus GG and Saccharomyces boulardii were two probiotics that showed promise. I went out on FMLA and returned to New York for three weeks to help my sister get mom home and ensure a consistent diet of gut rebuilding foods, including prebiotics."* | rfCDI |
| *"I was not going to get sick again, and I was willing to commit to healing by whatever method possible. After the 8 week period, I slowly reintroduced more foods, including building my gut bacteria back up with all kinds of fermentation strains: sourdough, sauerkraut, water kefir, 2 types of kombucha, etc. anything to get an abundance of probiotics in my body. Another thing I did was make sure to cut out any and all meat products from our diet that was not antibiotic-free."* | fCDI |

### Supplemental Figure S1

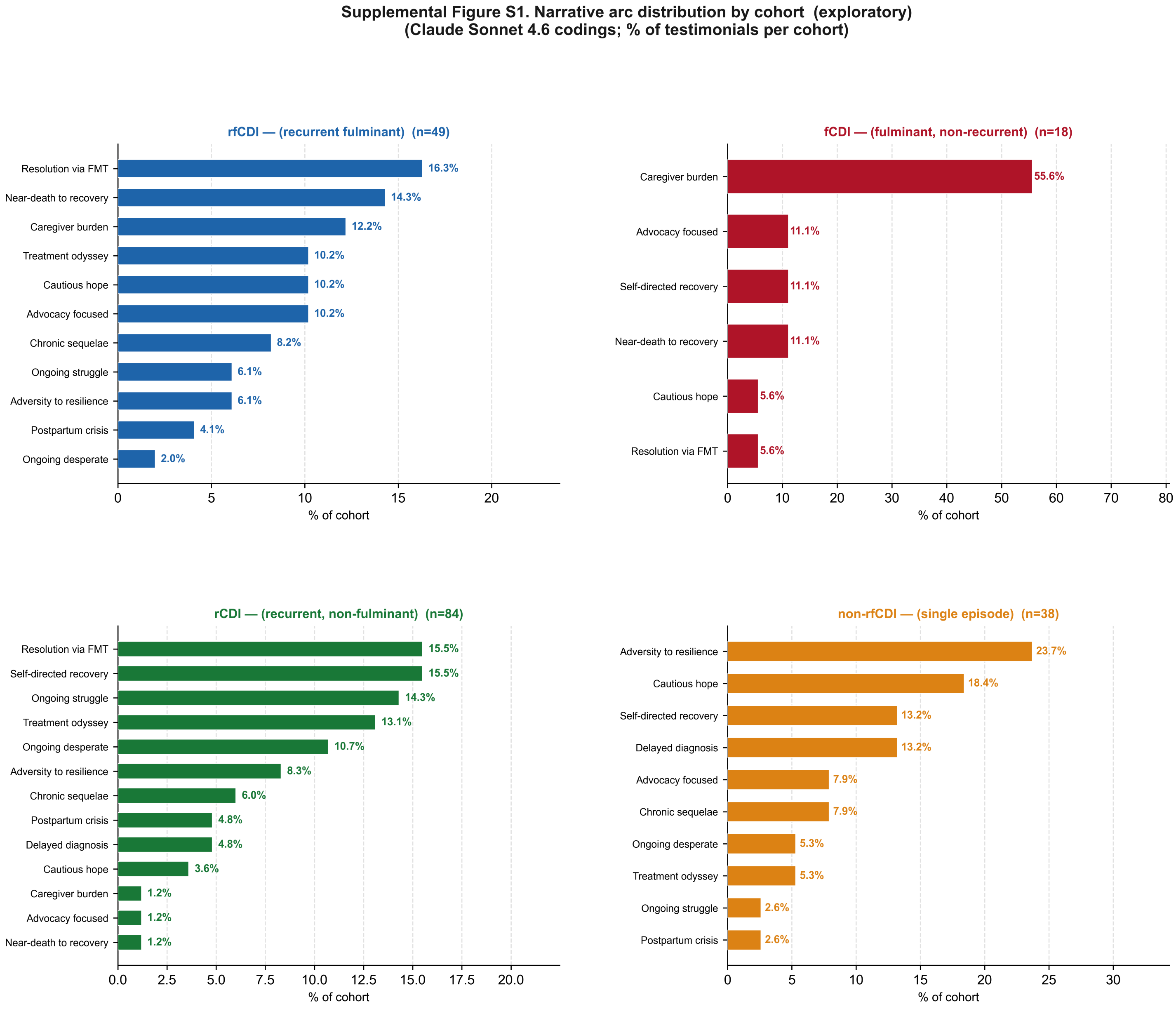

#### Supplemental Figure S2

**Supplemental Figure S2. Acquisition source and author characteristics**

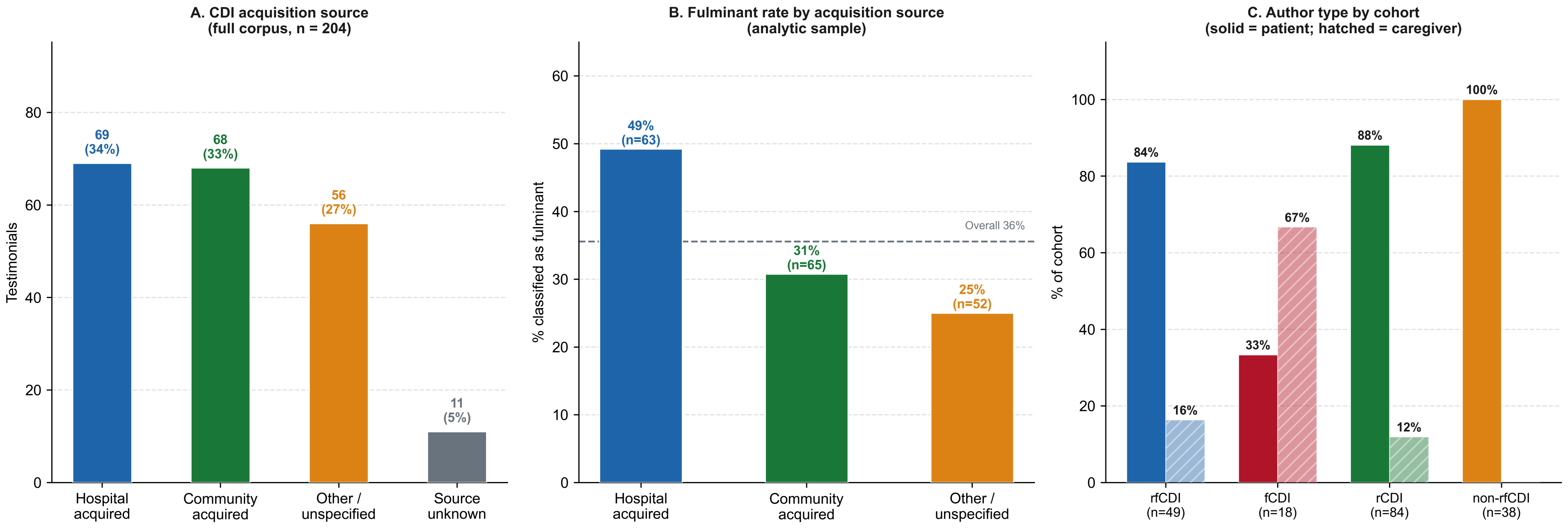

Supplemental Figure S3

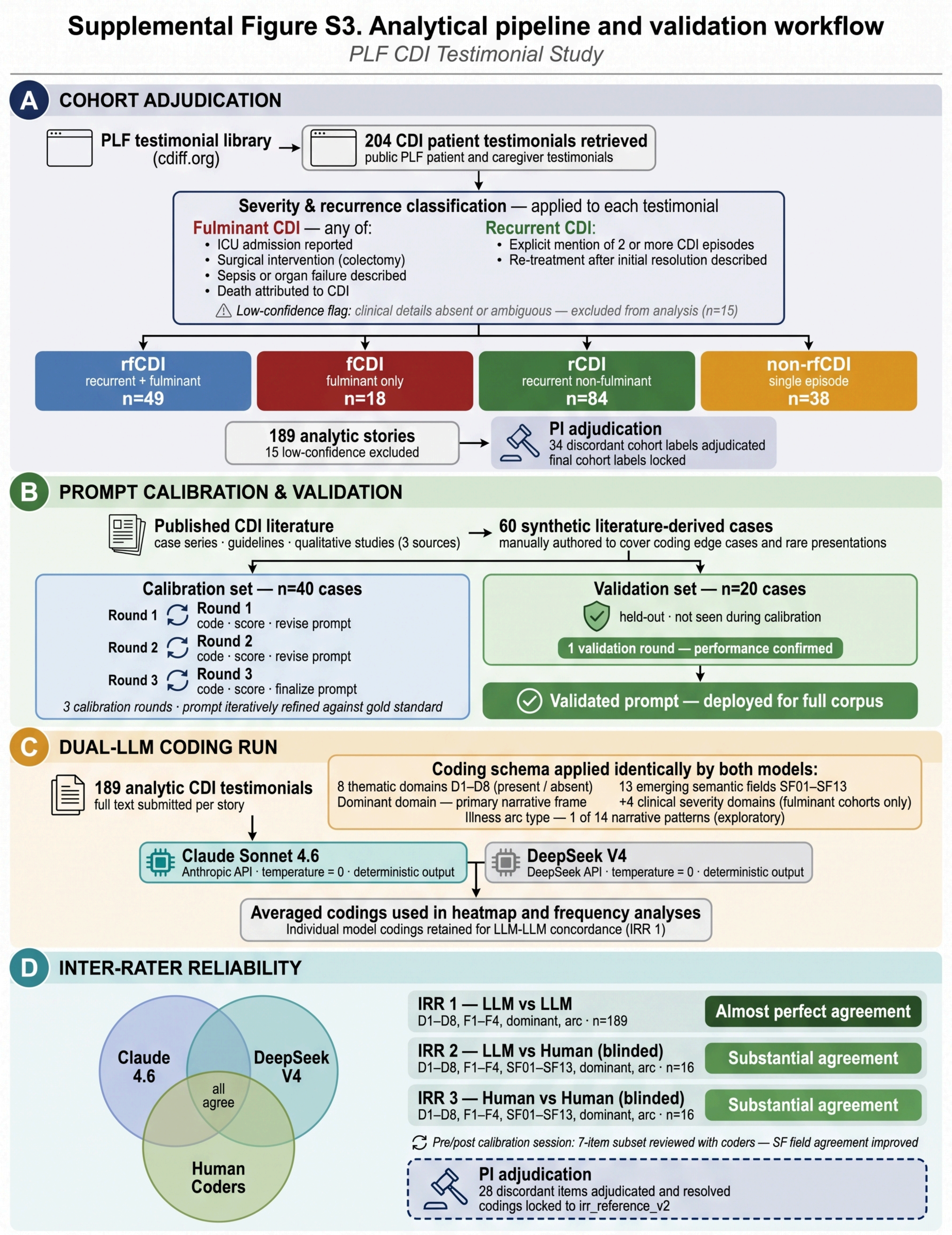


**CDI** = *Clostridioides difficile* infection. **rfCDI** = recurrent fulminant. **fCDI** = fulminant non-recurrent. **rCDI** = recurrent non-fulminant.
**SF** = emerging semantic field. **IRR** = inter-rater reliability. **LLM** = large language model.

Villafuerte-Galvez JA et al. 2026
